# Individuals with reported and novel *KDM5C* variants present with seizures, a feature recapitulated in a *Drosophila* model

**DOI:** 10.1101/2025.09.16.25335792

**Authors:** Bethany K. Terry, Amira Mahoney, Brian I. Lee, Julie Secombe

## Abstract

Variants in the chromatin regulator KDM5C cause a rare neurodevelopmental disorder (KDM5C-NDD) characterized by intellectual disability, seizures, and a broad range of systemic features. To better understand this disorder, more detailed and standardized information is required regarding the association between these genetic variants and cognitive and behavioral traits. Utilizing data obtained by the RARE-X KDM5C Data Collection Program, we analyzed survey and genetic data from 31 newly reported individuals. In addition to the expected neurodevelopmental challenges, participants frequently reported growth abnormalities, vision and digestive issues, behavioral concerns, and seizures in nearly half of cases. Meta-analyses of this data, combined with information from previously published cases, reaffirmed that seizures are a frequent feature in both male and female individuals with *KDM5C* variants, with over a third of individuals having had at least one seizure. Based on the prevalence of seizures in the RARE-X and published datasets, we next sought to explore the mechanisms underlying this behavior using the model organism *Drosophila* to develop robust quantitative assays of seizure-like behavior by modulating the expression of its single *Kdm5* gene. Loss of KDM5 specifically in neurons, but not glia, led to spontaneous and stimulus-induced seizures, underscoring a cell-intrinsic requirement for KDM5 in maintaining neuronal stability. Together, these human and fly studies highlight KDM5C as a critical regulator of nervous system function and demonstrate how patient-driven data collection and scalable model systems can be integrated. This work broadens our understanding of KDM5C-NDD and lays the foundation for future therapeutic discovery.

## I. Introduction

Neurodevelopmental disorders (NDDs) are a group of heterogeneous conditions that impact the development and function of the nervous system. Worldwide, it is estimated that over 316 million children live with NDDs.^1^ Individuals with NDDs present with a spectrum of symptoms, the presence and severity of which vary between individuals with different conditions and even amongst individuals with the same NDD. Genes encoding histone-modifying enzymes are increasingly recognized as a significant genetic cause of NDDs, including the Lysine Demethylase 5 (KDM5) family of histone demethylases.^2–4^

In mammals, the loss of KDM5A,^5,6^ KDM5B,^7–11^ or KDM5C function^12^ has been linked to atypical neurodevelopment, whereas KDM5D, located on the Y chromosome, has not. Of the three *KDM5* genes associated with NDDs, *KDM5C* has the largest number of reported variants, largely due to its X-linked inheritance, although many questions remain regarding how loss of KDM5C alters neuronal function. The disorder caused by pathogenic *KDM5C* variants has been known by multiple names (OMIM #300534), including “Claes-Jensen syndrome”, “Intellectual developmental disorder, X-linked, syndromic, Claes-Jensen Type (MRXSCJ)”, and “KDM5C-associated X-linked intellectual disability (KDM5C-associated XLID)”. Due to the variable expressivity of the traits observed, we have opted to use the more broadly inclusive term KDM5C-related neurodevelopmental disorder (KDM5C-related NDD, abbreviated as KDM5C-NDD). Like all KDM5 family proteins, the best-described role of KDM5C is its enzymatic role as a histone demethylase, working to remove di- and tri-methyl marks from Histone H3, at lysine 4 (H3K4) through the joint function of its Jumonji N (JmjN) and Jumonji C (JmjC) domains.^13^ H3K4me3 is a promoter-proximal mark associated with actively transcribed genes, indicating that variants affecting KDM5C activity likely impact brain function by altering gene expression. In addition to its canonical enzymatic activity, KDM5C and other KDM5 family proteins also impact gene expression through less characterized functions mediated by other regions of the protein.^14^ Both demethylase-dependent and independent functions of KDM5C are likely to be critical for its function in the development and functioning of the brain.^12^

Previous reports of individuals and families with *KDM5C* variants have noted a spectrum of neurological and other symptoms.^7,15–38, 39–63, 64,65^ The most prevalent features described include intellectual disability, developmental delay, fine and gross motor skill challenges, speech and language issues, short stature, epilepsy, and behavioral features like aggression, anxiety, attention deficit hyperactivity disorder (ADHD), and autism spectrum disorder (ASD). However, while the presence of some of these features, such as intellectual disability and short stature, is frequently reported in the literature, details related to traits such as seizures/epilepsy and impacts on organ systems outside the central nervous system (CNS) are often omitted. It is also notable that the literature tends to focus on describing the features of hemizygous males. Many studies overlook the traits associated with heterozygous females, although it should be noted that several recent articles have specifically focused on this issue.^21,39^ Therefore, additional and more comprehensive characterization of individuals with *KDM5C* variants is crucial to better understand this disorder and to determine whether any genotype-phenotype correlations can be identified.

Of the many clinical manifestations of KDM5C-NDD, seizures are one important but poorly understood feature, the presence of which can have dramatic quality-of-life consequences for the affected individual and their caregivers. Previous reports of larger cohorts of individuals with *KDM5C* variants in which seizures have been intentionally surveyed for have reported that between 13-20% of females and 33-53% of males present with seizures.^21,39,66^ However, seizure/epilepsy prevalence, manifestations such as the type, age of onset, frequency of episodes, and effective management strategies remain understudied and insufficiently characterized. Emphasizing this, several reports describe instances of KDM5C-NDD-associated seizures that cannot be controlled with conventional antiseizure medication (treatment-resistant; intractable).^24,39,64^ The prevalence of treatment-resistant epilepsy in KDM5C-NDD is not well understood due to lack of standardized reporting, however in the general population of individuals with epilepsy, it is estimated that 15% of children and 34% of adults fail to have their seizures well-controlled by medication.^67^ Beyond epilepsy associated with KDM5C-related NDD, KDM5A^6^- and KDM5B^9^-related NDDs are also associated with seizure susceptibility. It is therefore critically important to conduct additional research into the cellular and molecular mechanisms underlying KDM5-related seizures.

To study the mechanisms underlying KDM5-related NDDs that are difficult to investigate in humans, research has been conducted in *Drosophila* to better understand the roles that KDM5 proteins play.^68–72^ Unlike mammals, which possess four paralogous *KDM5* genes, *Drosophila* encode a single ortholog, known simply as KDM5, whose functions likely broadly reflect those of all mammalian KDM5 proteins.^12^ Work using this model is therefore relevant not only to KDM5C-NDD but also to KDM5A- and KDM5B-related NDD. *Drosophila* are well-established to display seizure-like behavior and have been used as a model to study the cell types and signaling pathways involved in seizure susceptibility for many years.^73,74^ Consistent with *Drosophila* being able to provide key insights into seizures caused by loss of *KDM5*-gene activity, we demonstrated that fly strains modeling specific pathogenic *KDM5C* missense variants exhibit an increased frequency of seizure-like activity in response to mechanical stress.^71^

In the current study, we extend our understanding of human KDM5C-NDD and expand our model of epilepsy in *Drosophila*. Using molecular and phenotypic data obtained from the rare-disease-centric data collection platform RARE-X, we identified 31 previously uncharacterized individuals from 29 families, which expands the number of unrelated cases of *KDM5C* variants in the scientific literature to 122 total. Phenotypic data was able to be obtained from 27 of the RARE-X-participating individuals, allowing us to characterize symptoms associated with KDM5C-NDD and to compare differences between sexes and between this cohort and previous reports. Through a meta-analysis of our data and published literature, we find that 82% of individuals present with intellectual disability, and that 35% of individuals present with seizures, with both features displaying sex-related differences between males and females. From our *Drosophila* studies, we find that we are reliably able to promote and characterize seizure-like behavior using heat-stress, mechanical-stress, and unprovoked paradigms filmed using a newly designed apparatus for holding fly vials during behavioral trials. Our tests revealed that loss of KDM5 in neurons, but not in glia, leads to seizure-like behavior. However, further investigation into the mushroom body as an epicenter of seizure-like behavior in our model revealed limited changes between control and knockdown groups, leaving us to propose future studies into the areas and neuron types that drive seizures.

## II. Results

### General Characteristics of 31 Newly Reported Individuals with *KDM5C* Variants

As a rare disorder, understanding the full spectrum of traits associated with KDM5C-NDD is an ongoing process. The classical description of KDM5C-NDD, first described by Stephan Claes and Lars Jensen, includes features such as short stature, intellectual disability, developmental delay, and motor challenges.^36,75^ While many reports focus primarily on these characteristics, more recent publications^21,39,66^ and anecdotal reports by members of the KDM5C-NDD community have suggested that the symptoms experienced by those with variants in *KDM5C* extend beyond the brain. Building on our evolving understanding of the heterogeneous nature of NDDs in general, we aimed to expand our knowledge of KDM5C-NDD by characterizing previously undescribed individuals with *KDM5C* variants.

To gain insight into the phenotypic spectrum of those with *KDM5C* variants, we mined data obtained by the 501(c)(3) non-profit organization RARE-X, a program of Global Genes. Through their KDM5C Data Collection Program, RARE-X recruited individuals with *KDM5C* variants or their caregivers (when applicable) to complete the Health and Development-Head-to-Toe and symptom-based surveys. Raw, de-identified data were accessed after approval from the Einstein Institutional Review Board (IRB) and the RARE-X Data Access Committee, following the completion of a Data Use Agreement. Using data provided in June 2025, we obtained information for 31 individuals with *KDM5C* variants and curatable genetic reports (Figure 1A). 28 unique variants were identified, with two unrelated individuals presenting with the same variant and three individuals from the same family presenting with the same variant (Table S1).

**Figure 1.**
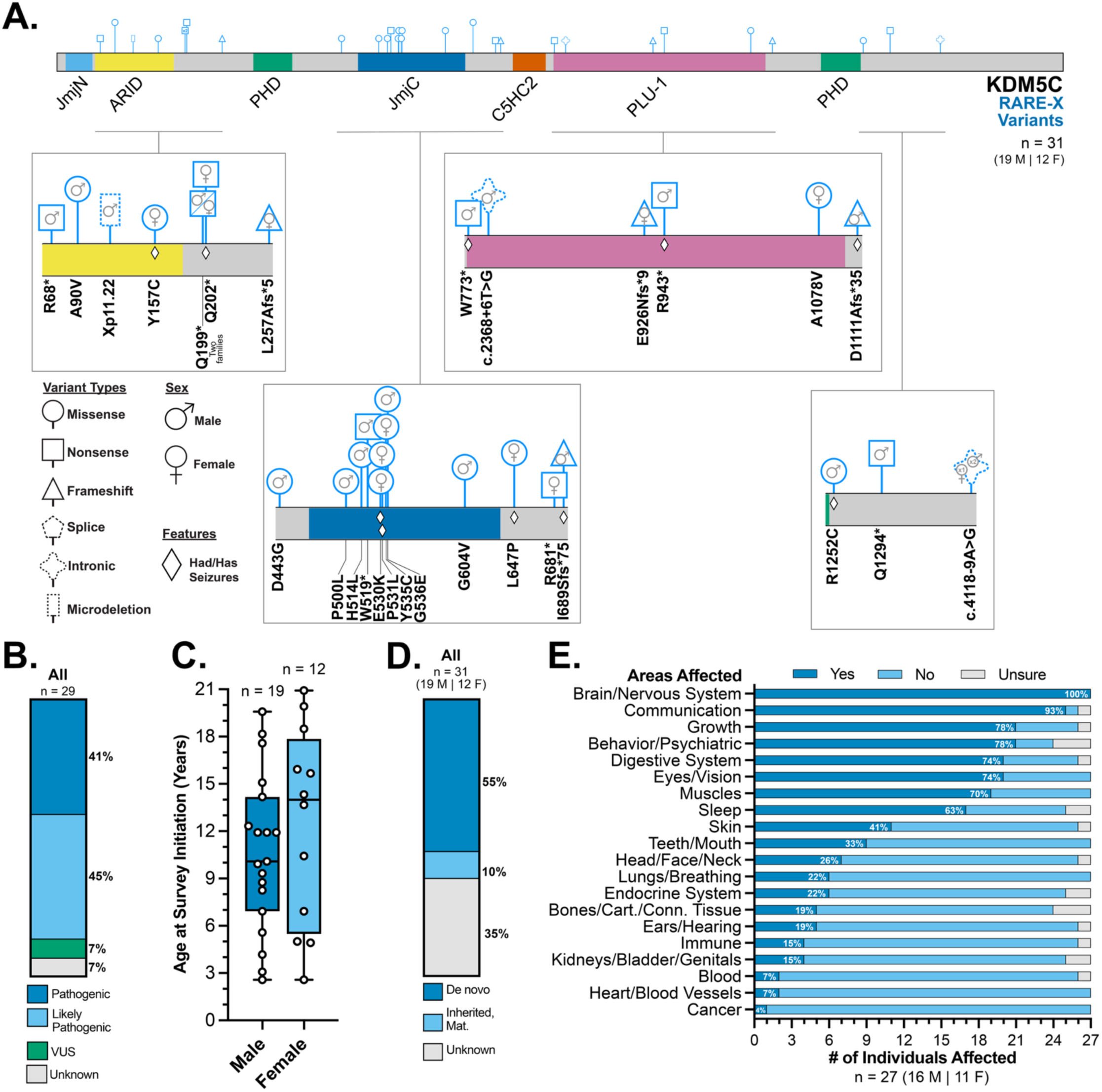
Demographic and phenotypic summary of 31 previously uncharacterized individuals with 28 unique KDM5C variants. (A) A schematic depicting the location, variant type, and sex of the individuals with variants in the protein KDM5C described in this study. Variants were reported to the non-profit organization RARE-X, a research program of Global Genes, and confirmed by the organization through assessment of uploaded genetic reports. Participants comprised of both males (M) and females (F) in this study. Variants in which seizures were reported in this study are annotated with a diamond. (C) Assessment of participants at the time of survey initiation showed that these individuals ranged in age from 2 years (31 months) old to 20 years (251 months) old. No significant difference was found in age between male and female participants (unpaired, two-tailed t-test, t = 0.8889, df = 29, P = 0.3814). (D) Review of available allele inheritance information revealed that the variants of this study’s subjects are primarily the result of de novo events, maternally inherited, or of unknown origin. (E) Analysis of completed Health and Development-Head-to-Toe surveys demonstrated that while all participants reported issues with their nervous system, a range of body systems were affected in individuals with *KDM5C* variants.

According to the provided information from the deidentified genetic reports, variants were detected by several means, which included whole exome sequencing, whole genome sequencing, multi-gene panel sequencing, and targeted familial variant testing. These alleles were distributed throughout KDM5C and were classified as pathogenic, likely pathogenic, variants of uncertain significance or were unclassified (Figure 1B). Of the 29 families represented, missense (13), nonsense (9), frame shift (4), intronic (2) variants were present, as well as 1 microdeletion (Figure 1A). A breakdown of participant information is provided in Table S1.

Both male (61%) and female (39%) participants were represented in this study. At the time of survey initiation, affected KDM5C-NDD individuals were between 2 years (31 months) and 20 years (251 months) old, with a median age of 10.6 years for male participants and 12.4 years for female participants (Figure 1C). For approximately one-third of reports, inheritance information was unavailable, possibly due to a lack of familial sequencing performed at the time of genetic report generation, adoption, or the submission of partial genetic reports (Figure 1D). Of the 31 participating individuals, over half were noted to have *de novo* variants, while around 10% reported maternally inherited alleles.

The first survey participants were asked to complete was a broad “Health and Development” survey, in which participants were asked about issues with different areas or body functions. Among the surveys beyond the general information collection form, the Health and Development survey had the highest participation, with a completion rate of 87%. Notably, in the Head-to-Toe subsection of the Health and Development survey (hereafter this subsection is referred to simply as the Head-to-Toe survey), all participants reported brain or nervous system issues (100%), and most (93%) reported challenges related to communication (Figure 1E). Other features that occurred in over half of the survey responders included phenotypes related to growth, behavior/psychiatric, the digestive system, eyes/vision, muscles, and sleep. For each of the other categories presented, at least one participant responded affirmatively to having had issues in that realm. Interestingly, despite reports linking KDM5C to some cancers, only one participant (4%) reported a malignancy, which they noted in a later survey was an unspecified neoplasm/neoplasia/tumor of the central nervous system.^76^

Comparing the frequency of features between sexes revealed no statistically significant difference using a Fisher’s Exact test. This could in part be due to our limited sample sizes, which decreases as surveys became more targeted. Therefore, we have chosen to not report additional inferential statistics for the rest of the RARE-X data set. Numerical differences existed between sexes (Supplemental Figure 1A-1B). For example, males never reported heart/blood vessel issues (0% of males, 18% of females) or cancer (0% of males, 9% of females). The largest difference between sexes was in the teeth/mouth category (25% of males, 45% of females).

As part of the overall Health and Development survey, participants were asked whether they had concerns about their development, when these concerns were first noted, and what specific concerns they had. Deviation from typical developmental milestones for males were noted between the ages of 3 and 30 months, with a median age of 8 months, and for females this occurred between the ages of 1 and 18 months, with a median age of 7.5 months (Figure 2A). Areas of developmental concern showed similar themes to those seen in the Head-to-Toe survey, with over half of respondents having reported challenges with motor development, muscle tone, and coordination (77-96%), communication, babbling/speaking (73-81%), and physical growth (65%) (Figure 2B). Other common developmental features included unusually intense interests in certain topics (54%), vision problems (46%), pointing/gesturing/imitating (42%), unusual responses to stimuli (38%), issues related to eating (35%), and self-injurious behavior (35%). Few features were dramatically different between sexes, with the greatest difference being a lack of pointing/gesturing/imitating (50% for males, 27% for females) (Supplemental Figures 1C-1D). In response to questions about bodily pain/discomfort, regression, and ambulation, most could walk without assistance (96%), although over a third reported having experienced regression (37%), with similar rates reported between males and females. 27% of participants stated they have bodily pain/discomfort (Figure 2C). This differed between sexes, as males (13%) and females (50%) responded yes to this question.

**Figure 2.**
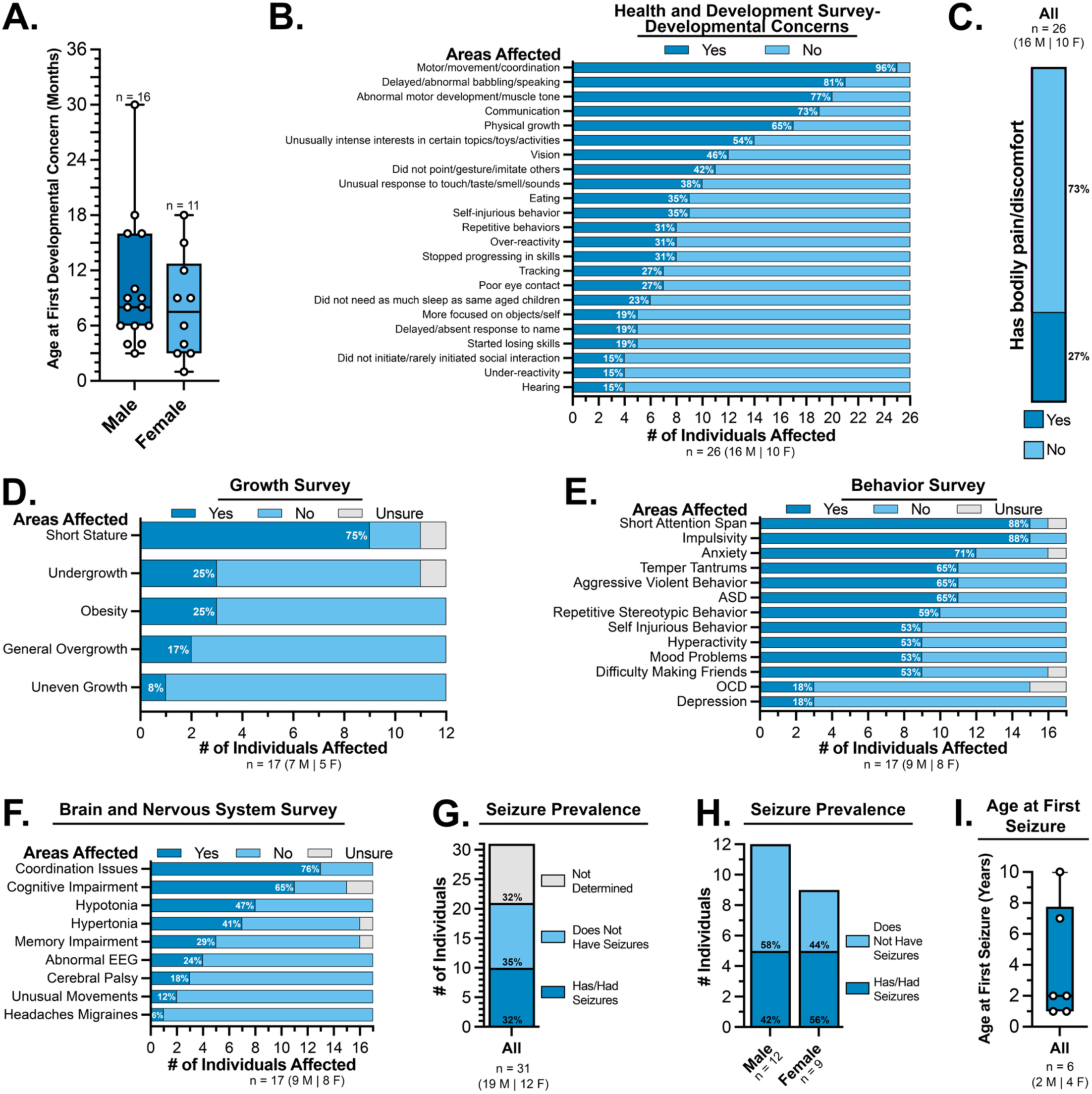
Individuals with KDM5C variants in the RARE-X cohort are frequently affected in the realms of development, growth, behavior, and nervous system function. (A) Assessment of completed Health and Development-Developmental Concerns surveys collected by RARE-X revealed that the individuals included in this study had their first developmental concern noted before the age of three, with some having concerns identified as early as one month in age. (B) Data from the Health and Development-Developmental Concerns surveys showed that motor and communication challenges were amongst the most prevalent developmental concerns reported by the individuals in this study. (C) Of those individuals with completed Health and Development surveys, more than a quarter of study individuals reported to have bodily pain or discomfort. (D) Short stature is the most frequently reported issue of the features noted by those who completed the level 2 (L2) Growth survey. (E) Analysis of the L2 Behavioral/Psychiatric survey results indicated that short attention span, impulsive behavior, and anxiety were some of the most commonly reported issues. Other features, including temper tantrums, aggressive behavior, and autism spectrum disorder, were noted in over half of the participants. Abbreviations used: Autism Spectrum Disorder (ASD), Obsessive Compulsive Disorder (OCD). (F) Evaluation of the features reported in the L2 Brain and Nervous System survey revealed that coordination issues and cognitive impairment were the most frequently reported features. Abbreviations used: Electroencephalography (EEG). (G) Similar percentages of individuals in our cohort have had a seizure before (32%) and have never had a seizure before (35%), while the remaining participants had no explicit seizure information available (32%). Seizure prevalence in our RARE-X cohort was calculated from data gathered from L2 Brain and Nervous System survey answers and information that RARE-X summarized from the genetic reports. (H) Seizure are a feature seen in many male and female participants. Seizure prevalence in our RARE-X cohort was calculated from data gathered from L2 Brain and Nervous System survey answers and information that RARE-X summarized from the genetic reports. (I) Pediatric Epilepsy Learning Healthcare System (PELHS) survey results showed the age at first seizure ranged between 1 and 10 years of age with a median age of 2.

### Study Participants Frequently Report Growth, Vision, Digestive, and Neurological Features

As expected, based on published descriptions of KDM5C-NDD, growth challenges were present in the majority of survey respondents (78% in the Head-to-Toe survey; Figure 1E). Results from the level 2 (L2) Growth Survey illustrated that 75% of KDM5C-NDD individuals exhibited short stature (71% of males and 80% of females) (Figure 2D, Supplemental Figure 1E-1F). Interestingly, females reported higher rates of multiple growth-related features, with the greatest difference observed in obesity, which was not seen in males but was reported for 60% of females (Supplemental Figure 1E-1F). The synthetic form of human growth hormone Somatropin was taken by two individuals for delayed bone age (1 individual) and delayed growth (1 individual).

74% of those in our cohort had eye or vision issues, which was also noted as an area of developmental concern (46%; Figures 1E and 2B). Responses from the L2 Vision survey highlighted that abnormal eye movement, such as strabismus or nystagmus, was present in the majority (91%) of individuals (100% of males, 80% of females) (Supplemental Figure 2A-C). Several other vision/eye-related issues were noted, however, the only other feature seen in more than one male and female participant was farsightedness (36% for all participants, 33% of males, 40% of females).

Digestive system challenges were also common in our participants (Head-to-Toe-74%; Figure 1E). Of the responders to the L2 Digestive System survey, the majority (83%) reported experiencing constipation, which was observed at the same rate among both sexes (83% of males and 83% of females) (Supplemental Figure 2D-F). Other features observed in at least a third of participants included defecation problems (42%), esophageal issues (33%), and feeding difficulties (33%). Intestinal, liver, and pancreatic issues were not reported by any participant. Medications taken for the management of constipation include osmotic laxatives such as polyethylene glycol (1 individual), lactulose (1 individual), and stimulant laxatives such as sennosides (1) individual.

78% of respondents reported behavioral or psychiatric concerns in the Head-to-Toe survey, with the L2 Behavior survey underscoring the presence of several features within our cohort (Figure 1E, Figure 2E, Supplemental Figure 2G-H). Over half of participants reported short attention span (88%), impulsivity (88%), anxiety (71%), temper tantrums (65%), aggressive behavior (65%), autism spectrum disorder (65%), repetitive stereotypic behavior (59%), self-injurious behavior (53%), hyperactivity (53%), mood problems (53%), and difficulty making friends (53%). Several of these features differed between males and females, including repetitive stereotypic behavior (78% of males compared to 38% of females), depression (0% of males and 38% of females), difficulty making friends (67% of males, 38% of females), and impulsivity (78% of males, 100% of females). Some individuals took medication for the management of behavioral/psychiatric concerns, which included sertraline (4 individuals) and risperidone (1 individual) for anxiety or depression, divalproex for aggression (1 individual), and methylphenidate (3 individuals), amantadine (1 individual), and guanfacine (1 individual) for ADHD or impulsivity.

From the Head-to-Toe survey, altered nervous system features were the most prevalent and best described of the features surveyed, with all respondents reporting changes (Figure 1E). The L2 Brain and Nervous System survey noted coordination concerns (76%), cognitive impairment (65%), hypotonia (47%), and hypertonia (41%; Figure 2F, Supplemental Figure 2I-J). Two of these traits showed sex differences: coordination, which was present in all females but only 56% of males, and unusual movements, which were not present in females but seen in 22% of males.

### Analyses Of Seizure Susceptibility Through Meta-analyses Integrating RARE-X And Published Data

A key focus of our analysis of the RARE-X data is seizures, for which information was pooled from multiple survey sources to determine the prevalence of this trait in our cohort. For 32% of participants, no seizure information was available. For the remainder of KDM5C-NDD individuals with seizure information, 48% have (or have had) seizures (Figure 2G). Seizures were observed in both male (42%) and female (56%) participants (Figure 2H). Of the six who completed the Pediatric Epilepsy Learning Healthcare System (PELHS) survey, the age at first seizure ranged from 1 to 10 years, with a median age of 2 years (Figure 2I). Several types of seizures were reported, including generalized tonic-clonic, motor, nonmotor, focal, and generalized seizures (Table S1). Some individuals presented with different combinations of these seizure types, while others were diagnosed with a singular type. For example, one individual reported having Doose Syndrome, also known as myoclonic atonic epilepsy, and another reported having Landau-Kleffner syndrome (Table S1). Several medications were used for seizure management, including the sodium channel blockers oxcarbazepine (2 individuals) and lamotrigine (brand name-Lamictal, 1 individual), the neuromodulator levetiracetam (brand name-Keppra; 2 individuals), and the GABA-A agonist midazolam (1 individual) for acute seizure control.

Although we have identified 31 previously unreported individuals with *KDM5C* variants and, at least in part, characterized the phenotypic features of these participants, we sought to integrate this information with the broader understanding of individuals with *KDM5C* variants. Following a literature search for papers relating to KDM5C-NDD, Claes-Jensen Syndrome, and *KDM5C* variants and neurodevelopment, we identified 101 families with *KDM5C* variants from 53 journal articles (Supplemental Figure 3A-B). Combining these data with our RARE-X cohort revealed a total of 122 unique variants from 130 families that we then conducted a meta-analysis on (Figure 3A). Within these combined 130 families, we identified a total of 269 individuals with *KDM5C* variants, comprising 153 males, 112 females, and 4 individuals for whom sex was not reported. Inheritance information was not available for a quarter of the identified individuals, due to a lack of reporting, adoption, or sequencing only of the proband (Figure 3B). Of the remaining cases, 17% possessed *de novo* variants, 47% inherited their variant maternally, 1% possessed paternally inherited alleles (two individuals from one family and one individual from an unrelated family), and 9% of cases stated the allele was inherited, but the parent of origin was not reported. A breakdown of the *KDM5C* variants that have been reported in the literature can be found in Table S2 and a simpler list of all of the KDM5C variants identified between this study and previous studies can be found in Table S3.

**Figure 3.**
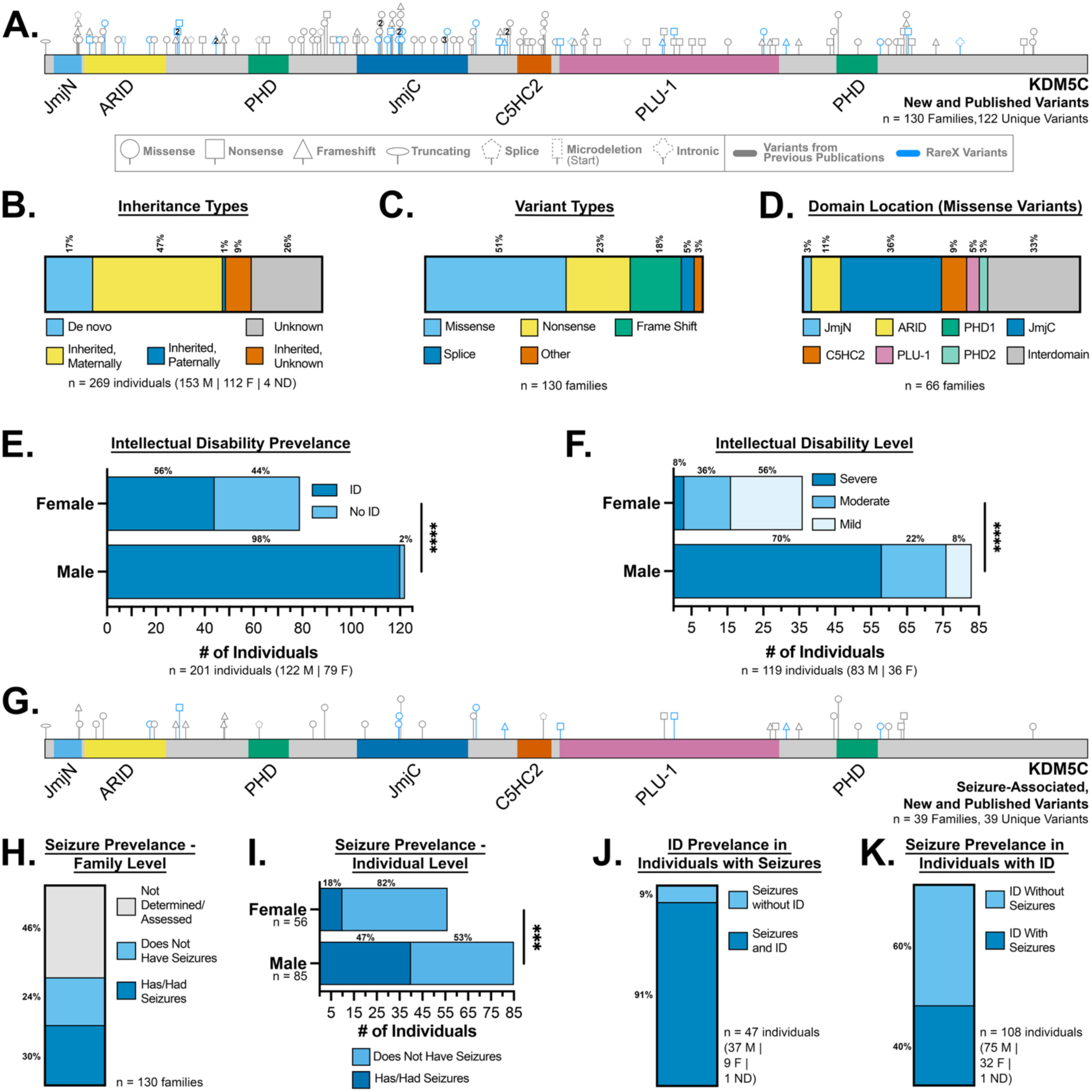
Assessment of the RARE-X cohort and previous reports reveal seizures and ID as common features in individuals with KDM5C variants. (A) A schematic illustrating the variant locations, variant types, and sexes of the individuals described in this study and reported in the literature. Variants in which there are more than one family with the same variant have a number in the center of the variant shape. (B) Analysis of the data collected by RARE-X and of information from previously published works indicates that maternal inheritance and *de novo* events are the most common means in which KDM5C variants are obtained. Paternal inheritance is a rare event in this population. Abbreviations used: Males (M), Females (F), Not Determined (ND). (C) Analysis of the data collected by RARE-X and of information from previously published works indicated that most reported variants are missense variants, followed by nonsense, frame shift, and splice variants. Other variants that occurred were intronic variants (2 instances), a microdeletion (1 instance), and a truncating (1 instance) variant. (D) Analysis of the data collected by RARE-X and of information from previously published works shows that KDM5C missense variants are found across the protein, within defined domains, and within interdomain regions. (E) Analysis of the data collected by RARE-X and of information from previously published works demonstrates that males with KDM5C variants are more likely to present with intellectual disability (ID) than females with KDM5C variants. (Fisher’s exact test, two-sided, P < 0.0001). ****P < 0.0001. (F) Analysis of the data collected by RARE-X and of information from previously published works indicates that males and females with KDM5C variants of those that present with ID show differences in the severity of ID present (Fisher’s exact test, P < 0.0001). ****P < 0.0001. (G) A schematic depicting the location, variant type, and sex of the individuals with KDM5C variants that have ever experienced seizures. Variants presented are from this study or previously published studies. (H) Analysis of the data collected by RARE-X and of information from previously published works indicates that while many reports do not explicitly mention seizures, 30% of families have at least one individual in the family that has had a seizure. Families and single individuals whose variant arose as a *de novo* event were counted as families. (I) Analysis of the data collected by RARE-X and of information from previously published works demonstrates that males with KDM5C variants are more likely to have seizures (or have had seizures) than females with KDM5C variants. (Fisher’s exact test, two-sided, P = 0.0006). ***P < 0.001. (J) Analysis of the data collected by RARE-X and of information from previously published works shows that the majority of individuals who have experienced seizures also present with ID. (K) Analysis of the data collected by RARE-X and of information from previously published works shows that over a third of individuals who have ID also present (or have presented with) seizures.

Variants were identified across the protein, with no obvious pattern in distribution (Figures 3A). Within the 130 families represented in this meta-analysis, about half of the variants were missense, 23% were nonsense variants, 18% were frame shift variants, 5% were splice variants, and 3% were other types of variants (which included a microdeletion, a truncating variant, and two intronic variants) (Figure 3C). To categorize these variants, analyses were done on a family level, rather than the total pool of individuals, to avoid duplication of variants (or inflation from families with different numbers of individuals represented).

Of the missense alleles identified, 33% were found in interdomain regions, while the remainder were within characterized motifs such as the JmjC domain, which had the largest number of variants (36%) (Figure 3D). This could be due to the fact that this region confers KDM5C’s canonical demethylase function, which is likely to play critical roles in pathogenesis, and that these individuals are potentially less likely to have their variants categorized as a variant of uncertain significance (VUS). Other domains with variants included the PLU-1 (5%), ARID (11%), C5HC2 (9%), JmjN (3%), and PHD2 (3%) domains. No missense variants were detected in the PHD1 domain. Nonsense variants were excluded from the domain location analysis due to the impact on multiple domains if a protein is produced, and the likelihood that these variants result in nonsense-mediated decay and no protein being produced. Likewise, microdeletions, frame shift variants, intronic, and splice variants were excluded from the domain location analysis due to the challenge in predicting the effects of these variants on overall protein structure.

Within the published literature, perhaps the most reported feature of KDM5C-NDD is intellectual disability (ID). Therefore, we sought to determine the prevalence of this feature in all 269 individuals and to investigate whether there was a sex-related difference, as previously reported.^39,66^ Of the 269 cases, information regarding ID was not available for 65 individuals. Of the 119 males and 77 females (201 annotated individuals) with ID information available, 82% were described as having ID (Supplemental Figure 3C). There was a significant difference between sexes in terms of ID occurrence, with 98% of males and 56% of females having documented ID (Figure 3E). Of those with reported ID severity, 51% presented with severe ID, 26% with moderate ID, and 23% with mild ID, although it is unclear whether the same criteria were used across all families (Supplemental Figure 3D). The percentage of each ID level varied significantly between sexes, with females generally reporting less severe cognitive impairment (Figure 3F). While 70% of males had severe ID, only 8% of females fell into this category. This trend is reversed for mild ID, with 8% of males and 56% of females, while moderate ID was more similar across the sexes, with 22% of males and 36% of females.

As seizures and epilepsy were a major feature that we noted in our RARE-X cohort, we next sought to assess seizure prevalence overall in the new and previously published cases. For this analysis, seizures were assessed at a family level, in addition to an individual level, to gain insights into potential genotype-phenotype connections, in addition to examining the penetrance of this trait. Any mention of epilepsy or seizures, even if the seizures were controlled or occurred once, was counted as the individual having had seizures. A simple list of the seizure-related variants identified can be found in Table S4. We identified 39 families with 39 unique variants associated with seizures distributed across the gene (Figure 3G). At a family level, seizure information was not described or assessed for 46% of cases (Figure 3H). Examination of only families with seizure information, 56% had seizures and 44% did not have seizures. On an individual level, of the 141 individuals for whom seizure presence could be determined, 35% of individuals had or have seizures (when sexes were pooled). When males and females were compared, a significant difference was noted between the sexes, with 47% of males having (or had) seizures, while 18% females having (or had) seizures (Figure 3I).

Where available, we also gathered information from the published literature about seizure types and medication taken (Table S2). Reported seizure types included generalized, generalized tonic-clonic, multifocal, focal, and absence seizures. Several individuals from one family had self-limited epilepsy, and one individual from a different family had sleep-related frontal hypermotor seizures. Previously published cases reported taking (or had previously taken) several medications for seizure control. Similar to the RARE-X data, these included sodium channel blockers such as levetiracetam (4 individuals), phenytoin (1 individual), and carbamazepine (3 individuals), and neuromodulators such as sodium valproate/valproic acid (5 individuals) and topiramate (1 individual). Several individuals reported their epilepsy to be intractable, however a lack of standard reporting relating to seizure presence and management made it difficult to identify the true proportion of individuals for whom no effective treatment could be found. Of those with seizures, 91% also had ID, suggesting that if you have seizures, it is likely that you will also present with ID (Figure 3J, Supplemental Figure 3E). However, for the reverse, the same does not hold true. Of those with ID, only 40% also had seizures, a rate similar to the overall population evaluated (37%) (Figure 3K, Supplemental Figure 3F).

### Developing A Robust System For Quantifying Seizure-Like Behavior In *Drosophila*

To gain insight into how *KDM5C* variants might lead to seizure predisposition and epilepsy, we turned to *Drosophila* as our animal model of choice. It should be noted that although the terms “seizure-like activity” or “seizure-like behavior” are the most accurate terms when describing this phenotypic behavior, hereafter, these terms are used interchangeably with the term “seizure” for simplicity. Previous work in *Drosophila* have utilized a variety of paradigms to bring about seizure-like activity-including through the use of mechanical, heat, cold, strobe light, and electrical stimulation to promote this behavior.^77–80^ We used mechanical and heat stimulation to promote seizure-like behavior, in addition to observing the behavior of flies without any induction method (spontaneous seizures) (Figure 4A). Bang-sensitive flies, which are more likely to seize following mechanical stimulation, undergo vortexing, a process that promotes seizure-like behavior by hyperstimulating sensory inputs.^77^ Heat-sensitive flies, in contrast, are more likely to seize following elevations in temperature to 37-42°C using a water bath or an incubator.^77,81^

**Figure 4.**
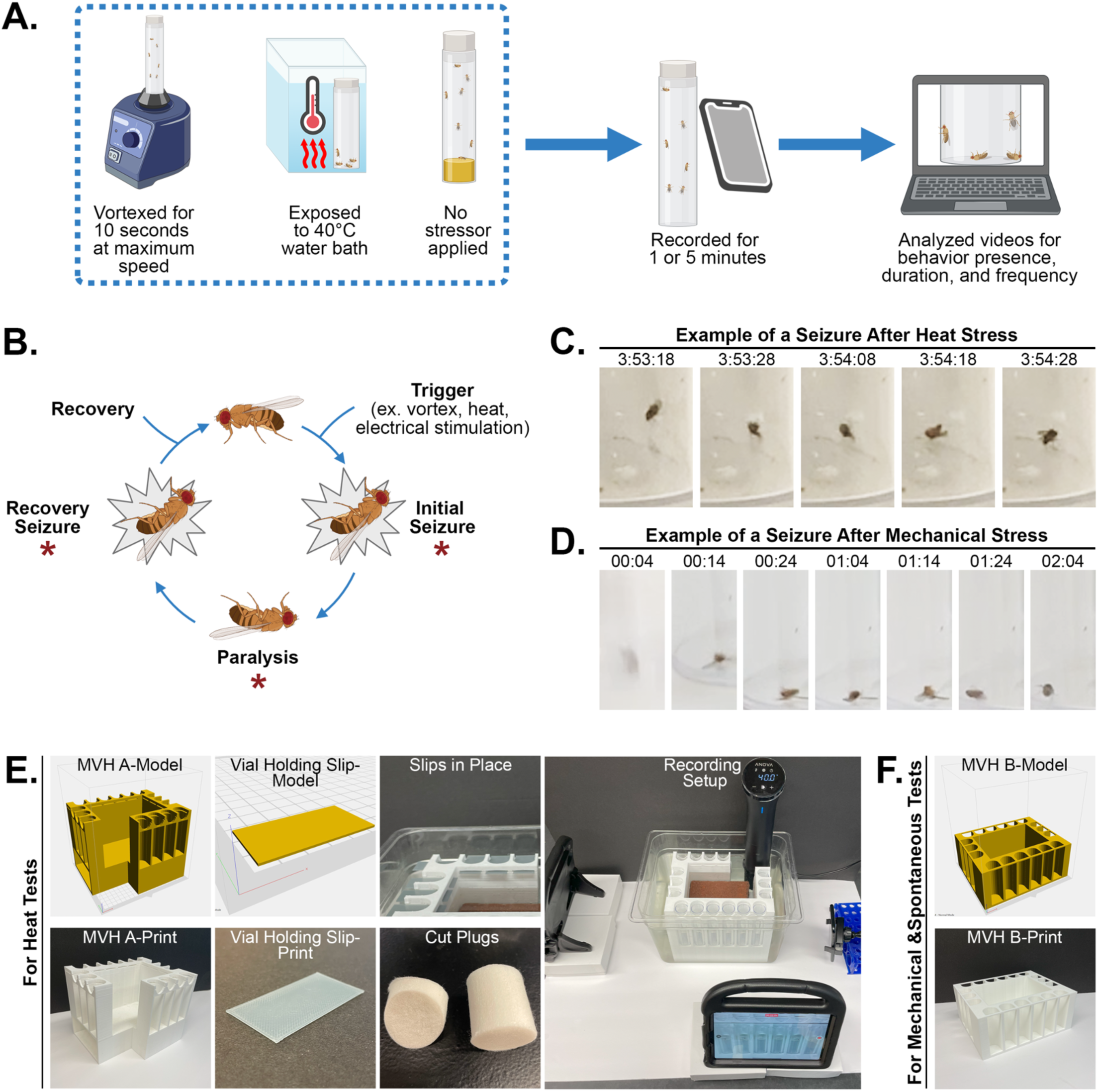
Experimental Setup and Presentation of Seizure-like activity in *Drosophila*. (A) An overview of the seizure induction methods, recording, and analysis procedure used in this study. Seizure-like activity was unprovoked or stimulated using heat or mechanical stress. Flies were recorded throughout this process for subsequent tracking in Adobe Premiere. (B) An illustration depicting the cycle of behavior during *Drosophila* seizure-like behavior. In this study we use the paralysis period as an indicator of seizure-like behavior and measure parameters such as loss of posture as an indicator of seizure-like behavior and measure parameters such as seizure duration, number of seizures, and time to first behavior. Loss of posture (a fly lying on their side or back) can take place with paralysis or can occur with shaking, wing flapping, or muscle contractions. Loss of posture occurs during the periods marked with asterisks. (C) An example of loss of posture following heat stress. (D) An example of loss of posture following mechanical stress. (E) Renderings and product images of the custom-designed multiple vial holder (MVH A) to allow the recording of multiple vials during heat seizure assays. (F) Rendering and product image of the custom-designed multiple vial holder (MVH B) for holding and recording vials during mechanical or spontaneous seizure assays.

For mechanical induction, flies were vortexed at maximum speed for 10 seconds, as previously described.^77^ For heat induction, flies in an empty vial were submerged in a 40°C water bath, ensuring the fly could not climb past the heated zone, similar to prior studies.^77^ For spontaneous seizure tests, flies were not given an additional stressor. For all conditions, flies were recorded, and videos were scored for the presence, duration, and frequency of seizure-like behavior (Figure 4A). To quantify seizure-like behavior, flies were scored as seizing if they exhibited a loss of posture, characterized by lying on their side or back with their legs in the air for at least one second. This is a behavior that is readily observable (Figure 4B-D). Loss of posture is an established component of *Drosophila* seizure-like behavior and can be accompanied by immobilization (paralysis) or movement that includes wing flapping, leg shaking, and abdominal muscle contractions (as seen in initial and recovery seizures) (Figure 4B).^73,74,82^ For heat induction, these seizure-like behaviors tended to occur after a minute or more of heat exposure, while for mechanical induction, these behaviors were present early in the recordings, sometimes occurring in the first frame of the video recording after vortexing.

While mechanical and heat stimulation have been successfully utilized for several years, there is no standardized method, with observation time varying and seizure behaviors often being scored in real-time. To ensure that every seizure-like behavior was accurately identified, flies were recorded, and behaviors were quantified through detailed analyses of the videos. In addition to providing accurate seizure quantification, this system allowed us to simultaneously record multiple behavioral trials. However, after running several trials, we noted several challenges with our behavioral setup for all three assays. Key among these were 1) we needed to develop a sturdy apparatus to hold our vials in a consistent location to keep vials in frame of the camera; 2) we needed our vials to be located in front of a solid, consistent background to make visual tracking of the fly easier; and 3) we needed a apparatus that could hold our fly vials, which are primarily filled with air, beneath the surface of water bath when testing flies in our heat paradigm.

To address these challenges, we designed two custom 3D-printable models to hold multiple standard, narrow-sized *Drosophila* vials simultaneously (Figure 4E-4F). The first multiple vial holder (MVH A) was designed for use in a water bath. For our heat seizure tests, we repurposed an immersion circulator designed for sous vide cooking to provide a consistent temperature throughout the water bath. This system additionally allows us to record the flies throughout their time exposed to heat by immersing the immersion circulator in a clear plastic container. Our MVH design A accommodates up to 16 vials simultaneously, while also providing space for the immersion circulator. To prevent the plastic apparatus from floating, space was left open in the center and at the bottom of the MVH for weights to be added (in our case, bricks). To prevent the vials themselves from floating, slots were added to the MVH to keep them underwater. All four sides of the apparatus can be filmed, depending on the number of recording devices available. To ensure that the fly was always in view, plugs were cut at an angle to limit recording blind spots. For non-heat-induced seizure analyses, a second design was created that can hold up to 18 vials at once (MVH B, Figure 4F). The MVHs can be printed inexpensively using the supplied files if a 3D printer is already accessible. Recording in our new 3D-printed apparatus enabled flies to be seen clearly, even when recording multiple vials simultaneously (Figure 4C-D).

### *Drosophila* KDM5 is Required in Neurons to Prevent Spontaneous and Induced Seizure-like Behavior

Both glial and neuronal dysfunction can contribute to seizures in humans and other mammals, as the roles of glial cell types, such as astrocytes, microglia, and oligodendrocytes, are significant in the regulation of neuronal activity.^56,83–85^ Neuronal and glial dysfunction can similarly contribute to seizures in *Drosophila,* with perturbation of different proteins exclusively in either cell type having been linked to seizures.^81,86–89^ Previous work from our lab showed that missense variants in *Drosophila* that model KDM5C-NDD alleles displayed increased susceptibility to seizures in response to mechanical stress. However, because these strains are whole-body mutants, the cell types involved in driving these seizures remained undetermined.

To determine whether KDM5 was required in neurons, we used the UAS/GAL4 system to drive the expression of a short hairpin RNA targeting *Kdm5* in neurons only, using the Elav-GAL4 driver. To confirm knockdown of *Kdm5*, we used immunohistochemistry on adult brains, labeling neurons using an antibody against Elav and labeling KDM5 using an antibody against the hemagglutinin (HA) tag that we knocked into the C-terminus of the *Kdm5* gene.^69^ In the brains of control animals, KDM5 was broadly expressed in neurons, and this was significantly reduced in *elav*>*shKdm5* flies (Figure 5A-5B). Despite the reduced expression of KDM5 in neurons, the brains of knockdown animals appeared indistinguishable from those of wild-type animals in terms of size and overall number of neurons. Whole-body perturbation of KDM5 or reduction of KDM5 in precursor cells of a key learning and memory structure, the mushroom body, can lead to axonal growth and guidance defects. To determine if neuronal knockdown of *Kdm5* caused similar defects, we examined the axonal morphology of the α/β lobe in the mushroom body by detecting the cell adhesion molecule Fasciclin II (FasII).^69,71^ 46% of adult brains from *elav*>*shKdm5* showed growth and guidance defects in the α/β lobe, which were not observed in control animals (Figure 5C-5D).

**Figure 5.**
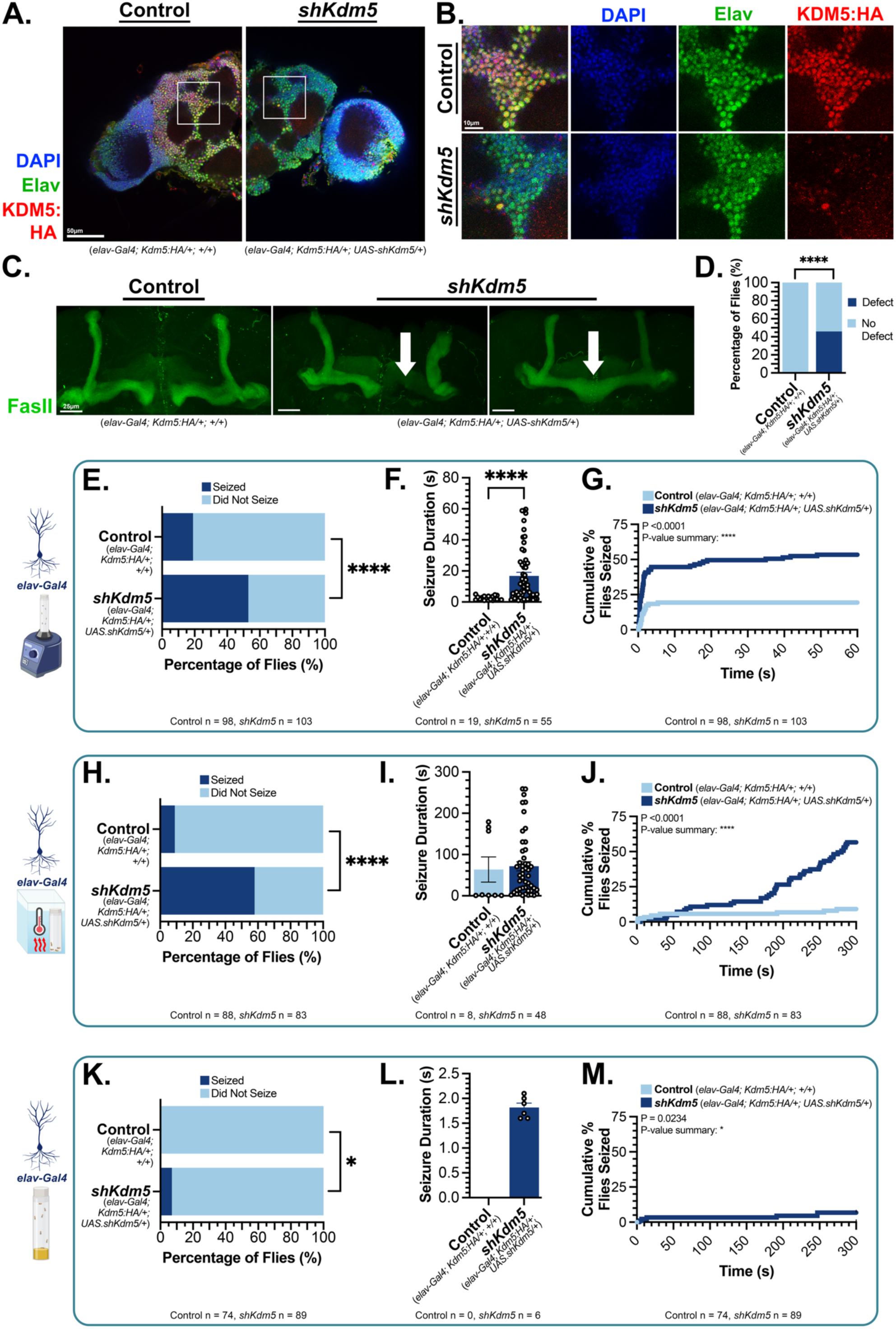
KDM5 is expressed in neurons and its loss causes seizure-like behavior and mushroom body defects. (A) Central brain region from adults from *elav*>+ control (*elav-Gal4*; *Kdm5:HA/+*; *+/+*; left panel) and *elav*>*shKdm5* knockdown (*elav-Gal4*; *Kdm5:HA/+*; *UAS-shKdm5/+*; right panel) in a strain that has endogenous KDM5 C-terminally tagged with a hemagglutinin (HA) epitope. Anti-HA detects endogenous KDM5:HA, which is observed in most neurons detected by anti-Elav. KDM5:HA is significantly reduced in Elav positive cells in *elav*>*shKdm5* animals. Maximum projection images were generated from immunostained adult fly brains (collected ≤7 days post-eclosion). Scale = 50 µm. (B) Magnified images from the boxed regions in panel A showing reduced KDM5:HA only in knockdown animals. Scale = 10 µm. (C) *elav*>*shKdm5* and not *elav*>+ animals show growth and guidance defects (white arrows) in mushroom body neurons detected using anti-FasII. Scale = 25 µm. (D) Quantification of growth and guidance defects presented in panel E. Control n = 40, *shKdm5* n = 67. Statistical analysis was done using a Fisher’s Exact test (two-sided, P < 0.0001). ****P < 0.0001. (E-G) *elav*>*shKdm5* flies seized significantly more frequently and for longer durations than controls in response to mechanical stress. Statistical analysis for panel E was done using a Fisher’s exact test (two-sided, P < 0.0001). Statistical analysis for panel F was done using a Mann-Whitney test (two-tailed, P < 0.0001). Statistical analysis for panel G was done using a Log-rank (Mantel-Cox) test (P < 0.0001). ****P < 0.0001. (H-J) *elav*>*shKdm5* flies seized significantly more frequently and for longer durations than controls in response to heat stress. Statistical analysis for panel I was done using a Fisher’s exact test (two-sided, P < 0.0001). Statistical analysis for panel J was done using a Mann-Whitney test (two-tailed, p = 0.1667). Statistical analysis for panel K was done using a Log-rank (Mantel-Cox) test (p<0.0001). ****P < 0.0001. (K-M) *elav*>*shKdm5* flies seized significantly more frequently, but no longer in duration, than controls in response to no stimulus. Statistical analysis for panel K was done using a Fisher’s exact test (two-sided, P = 0.0323). Statistical analysis for panel L was not able to be undertaken, given that no control flies seized spontaneously. Statistical analysis for panel G was done using a Log-rank (Mantel-Cox) test (P = 0.0234). *P < 0.05.

To determine if our flies were bang-sensitive, control and *elav*>*shKdm5* were subjected to the vortex mechanical seizure assay. A significantly larger proportion of knockdown flies exhibited seizure-like behavior (53%) when compared to controls (19%) (Figure 5E). Not only did *shKdm5* flies seize more frequently, but they also exhibited longer seizure duration periods (with a mean of 2.5 seconds) than controls (with a mean of 16.8 seconds) (Figure 5F). Quantifying the cumulative proportion of seizure-like activity over one minute revealed that more *shKdm5* flies exhibited seizure-like activity and that this occurred more quickly after vortexing than control flies (Figure 5G). A one-minute timeframe was utilized given the quick responses of the flies following the mechanical stimulus. Although *Kdm5* knockdown flies were more likely to have a seizure, the number of events per fly was not different from controls, with most flies having only one seizure (Supplemental Figure 4A-C).

In addition to being sensitive to mechanical stress, *elav*>*shKdm5* flies also showed a significant susceptibility to heat-induced seizures (Figure 5H-J). During heat stress, as in our mechanical stress paradigm, both control and knockdown displayed seizure-like behavior, although *shKdm5* flies showed a significantly increased overall seizure susceptibility (58%) when compared to controls (9%) (Figure 5H). Quantifying the seizure-like events over the five-minute window of observation, the differences between groups began to become apparent after the first minute of recording, with the total number of flies having seized continuing to increase over time (Figure 5J). However, in contrast to the seizure duration periods exhibited following mechanical stress, no significant difference in duration was noted between the control and *shKdm5* groups (Figure 5I). The number of seizure events also did not significantly differ between groups (Supplemental Figure 4D-F).

In addition to characterization of seizure-like behavior following application of a stressor, we sought to assess this behavior at baseline without the application of a stressor. This was done by placing vials into the fly holding apparatus described in Figure 4. Significantly more *shKdm5* flies seized (6.74%) than controls (none of which seized during these tests) (Figure 5K and 5M). Seizure duration and number of seizures, however, could not be compared between groups, as no flies exhibited seizure-like behavior in the control group (Figure 5L Supplemental Figure 4G-I)

Because individuals with *KDM5C* variants exhibit sex-related differences in seizure prevalence, we examined whether male and female flies showed similar increases in seizure susceptibility. For mechanical and heat stress conditions, male and female flies showed similar rates of seizure susceptibility (Supplemental Figure 5A, 5C, 5E, and 5G). Seizure duration showed similar increases for mechanical seizures between sexes (Supplemental Figure 5B and 5D), while, as in the pooled analyses, no difference was detected in seizure duration in the heat condition (Supplemental Figure 5F and 5H). For the spontaneous seizure condition, when separated into female and male groups, no significant difference was detected between control and knockdown groups, perhaps due in part to the limited number of flies that seized (Supplemental Figure 5I-L).

### KDM5 Activity Is Not Required In Glia Or Mushroom Body Neurons To Suppress Seizures

As described earlier in this manuscript, dysfunction of neurons and glia can contribute to the pathophysiology of epilepsy. Previous work in *Drosophila* has shown that both overexpression and knockdown of different genes, solely in glia and not in neurons, can promote heat-related, mechanical-related, and spontaneous seizure-like behavior, indicating that manipulation of glia is sufficient to cause neuronal dysfunction and ultimately seizure-like behavior.^81,86–88^ To test whether glia contribute to the seizure-like behavior in our model, we reduced KDM5 levels by using the pan-glial driver Repo-GAL4 to knock down *Kdm5* using the same short hairpin transgene.

KDM5 is expressed in the majority of glial cells examined, and *repo*>*shKdm5* efficiently reduced levels of KDM5 in Repo-positive cells of the adult brain (Figure 6A-6B). As glia can be a driver of mushroom body defects, we also probed our *repo*>*shKdm5* knockdown brains for morphological defects in the ⍺/β lobes.^90–92^ Anti-FasII staining revealed that only a small portion of the *repo*>*shKdm5* knockdown flies showed growth defects (6.5%) (Figure 6C-6D). While this was significantly increased compared to control flies, which did not show any defects, it was far lower than the defects seen in *elav>shKdm5* flies (46%). *repo>shKdm5* flies were not, however, more likely to exhibit seizure-like behavior than controls without an applied stressor or following heat or mechanical stress (Figure 6E-6G). Nor were changes in seizure duration or the cumulative percentage of flies seized over time different in animals with reduced KDM5 in glia (Supplemental Figure 6A-6F). Finally, separating flies into female and male-specific comparisons revealed no significant differences between groups, except for seizure duration for the female flies undergoing the vortex assay (Supplemental Figure 7A-7L). *shKdm5* flies in this condition, however, showed an overall lower susceptibility to seize. Reduced levels of KDM5 in glial cells is therefore not sufficient to drive seizure-like behavior in flies.

**Figure 6.**
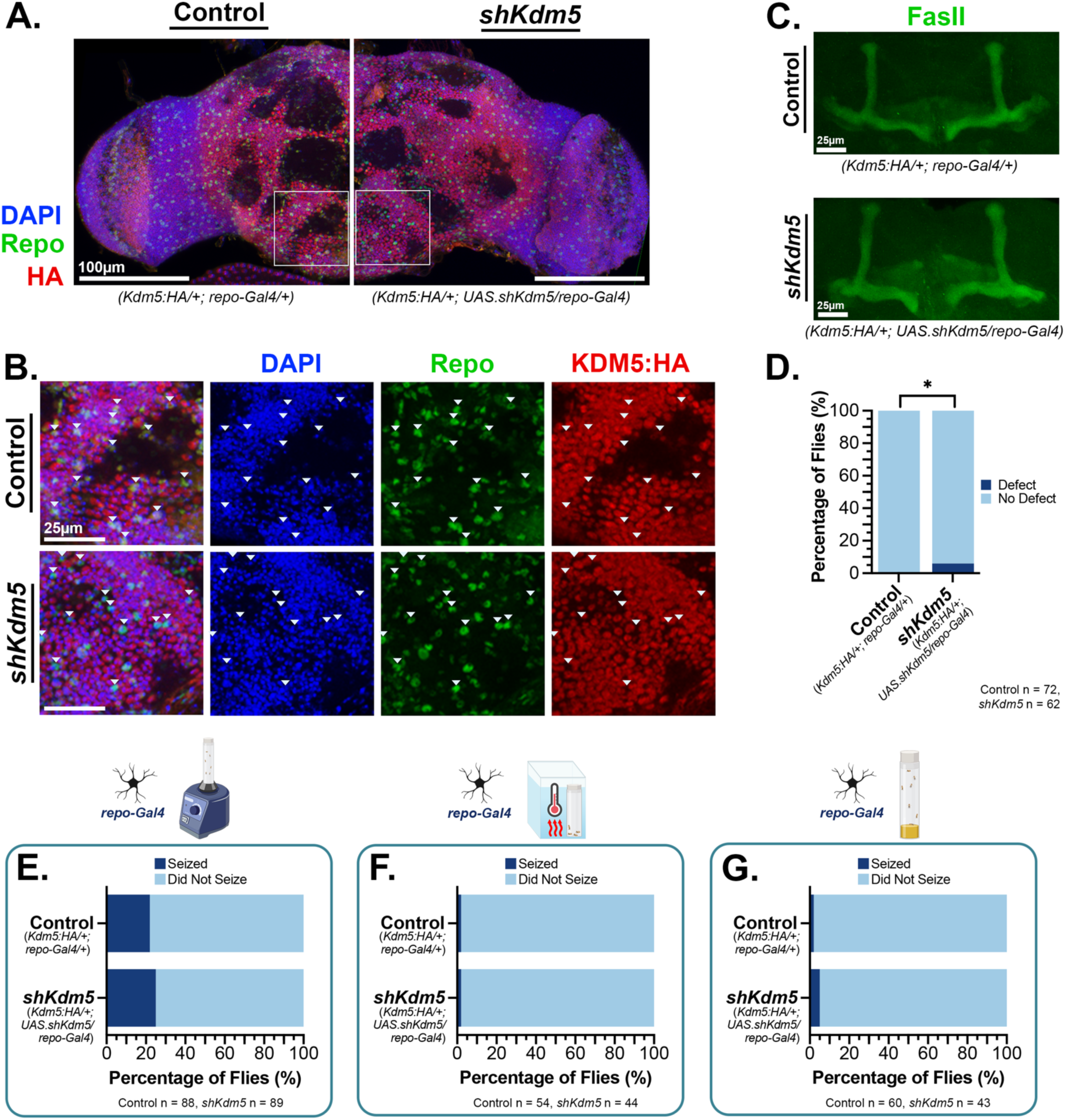
KDM5 is expressed in glia and its loss causes limited mushroom body defects, but not seizures. (A) Endogenously tagged KDM5:HA is found in many glia (Repo+) of the central brain in *repo>+* controls (*Kdm5:HA/+; repo-Gal4/+*) animals and is greatly reduced upon *Kdm5* knockdown in *repo>shKdm5* (*Kdm5:HA/+; UAS.shKdm5/repo-Gal4*) animals. Maximum projection images were generated from immunostained adult fly brains (collected ≤7 days post-eclosion). Scale = 100 µm. (B) Magnified images from the boxed regions in panel A. Arrowheads indicate Repo positive cells. Scale = 25 µm. (C) FasII immunostaining shows no structural changes in alpha and beta lobes of the adult mushroom body in the majority of flies following *Kdm5* knockdown in glia in *repo>shKdm5* (*Kdm5:HA/+; UAS.shKdm5/repo-Gal4*) animals. (C) Reduction of KDM5 levels in glia affects mushroom body structure in 6% of animals studied. Statistical analysis for was done using a Fisher’s exact test (two-sided, P = 0.0434). *P < 0.05. (E) *repo*>*shKdm5* flies showed no difference in seizure susceptibility or duration in response to mechanical stress. Statistical analysis was done using a Fisher’s exact test (two-sided, P = 0.7265). (F) *repo*>*shKdm5* flies showed no difference in seizure susceptibility or duration in response to heat stress. Statistical analysis was done using a Fisher’s exact test (two-sided, P > 0.9999). (G) *repo*>*shKdm5* flies showed no difference in seizure susceptibility or duration when observed without an induced trigger. Statistical analysis was done using a Fisher’s exact test (two-sided, P > 0.5696).

Previous work has established that cells of the mushroom body can be an origin for seizure-like behavior in *Drosophila*.^93,94^ Based on our finding that Elav-GAL4, but not Repo-GAL4, driven knockdown of KDM5 caused both seizures and neuromorphological changes in the mushroom body, we specifically assessed the role of these cells. To test whether the mushroom body could be an important area for KDM5-mediated seizure generation, we knocked down *Kdm5* using the well-characterized OK107-GAL4 driver. As we have shown previously, OK107-GAL4 driven knockdown of *Kdm5* produces animals with striking mushroom body defects in the adult brain (Figure 7A).^69^ Despite structural defects in the mushroom body, no statistically significant difference in mechanical stress or heat stress was observed (Figure 7B-7J, Supplemental Figure 8A-8L). For the spontaneous seizure condition, the overall susceptibility trended towards significance (P = 0.0577), and the assessment of susceptibility over time using a survival curve revealed a significant difference between groups (P = 0.0281). None of the control flies in this condition displayed seizure-like behavior, whereas we observed seizure-like behavior in our knockdown flies. Together, this data suggests that while mushroom body neurons may contribute slightly to the development of this behavior, the mushroom body is not an epicenter of seizure-like behavior in our KDM5-deficient flies.

**Figure 7.**
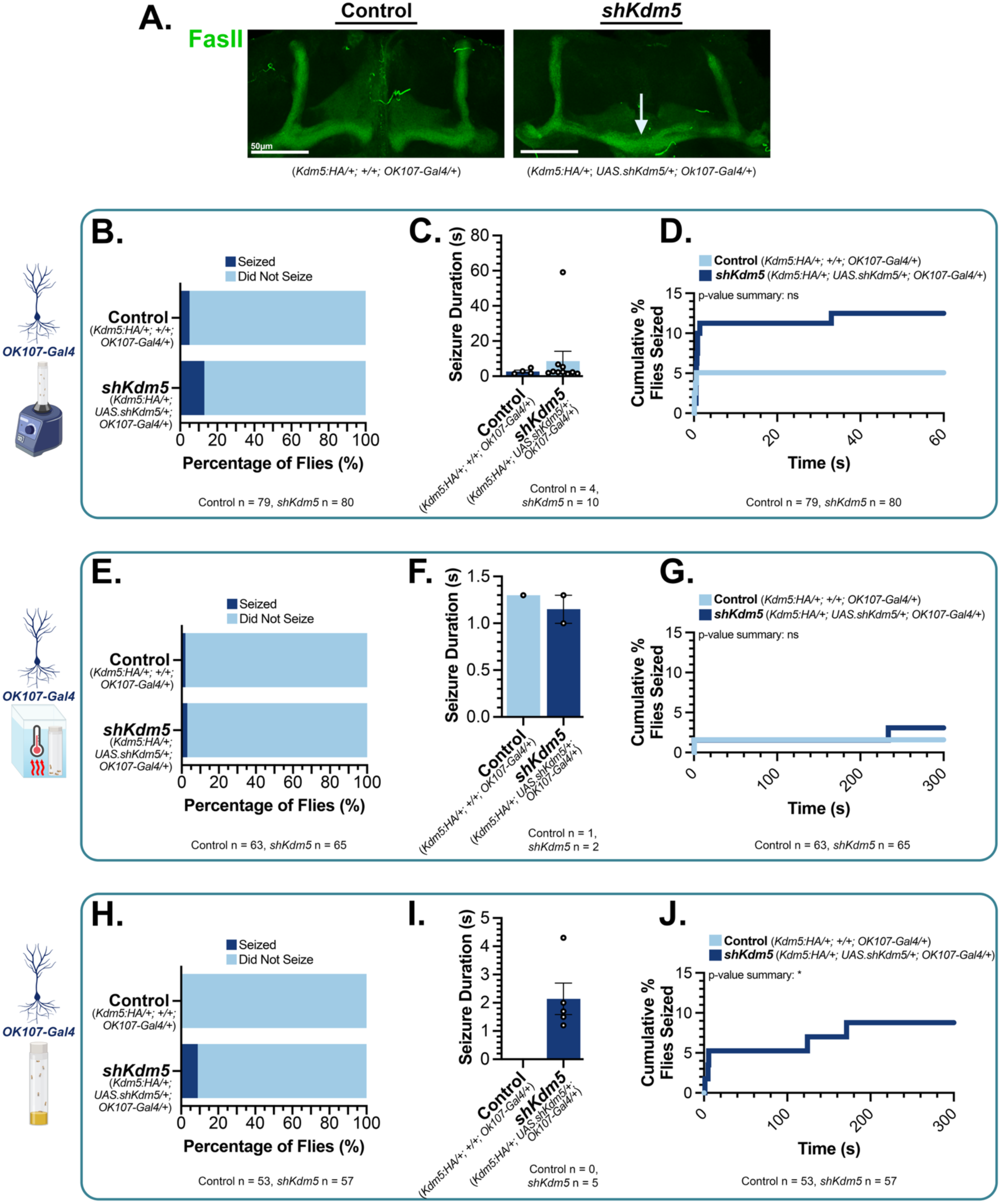
KDM5 reduction in mushroom body neurons promotes mushroom body defects, but not seizures. (A) Anti-FasII staining shows that reduction of KDM5 levels in mushroom body neurons promotes growth and guidance defects (white arrows), which are not seen in controls. Control genotype is *OK107*>+ (*Kdm5:HA/+; UAS.shKdm5/+; OK107-Gal4/+*) and knockdown *OK107*>*shKdm5* genotype is (*Kdm5:HA/+; UAS.shKdm5/+; Ok107-Gal4/+*). Scale = 50 µm. (B-D) *OK107*>*shKDM5* and *OK107*>+ flies showed similar seizure rates and durations following mechanical seizure induction. Statistical analysis for Panel B was done using a Fisher’s Exact test (two-sided, P = 0.1600). Statistical analysis for panel C was done using a Mann-Whitney test (two-tailed, P = 0.7103). Statistical analysis for panel D was done using a Log-rank (Mantel-Cox) test (P = 0.1092). (E-G) *OK107>shKDM5* and *OK107*>+ flies seized at indistinguishable rates and durations following heat seizure induction. Statistical analysis for Panel E was done using a Fisher’s Exact test (two-sided, P>0.9999). Statistical analysis for panel F was not able to be completed because of the limited number of flies that seized. Statistical analysis for panel G was done using a Log-rank (Mantel-Cox) test (P = 0.5791). (H-J) *OK107*>*shKDM5* and *OK107*>+ flies seized at similar rates and durations when observed for spontaneous seizures. Statistical analysis for Panel H was done using a Fisher’s Exact test (two-sided, P = 0.0577). Statistical analysis for panel I was not able to be completed because of the limited number of flies that seized.

## III. Discussion

In this study, we characterized 31 individuals from 29 families with 28 unique variants. While several other reports have provided sizable cohorts of individuals with *KDM5C* variants (notably Leonardi et al., 2022; Carmignac et al., 2020; Jensen et al., 2005; and Tzschach et al., 2006), our study has provided the largest cohort of families described to date. Through our analysis of body-wide symptoms, we have noted several features observed in the majority of participants, including short stature, coordination issues, cognitive impairment, short attention span, impulsivity, anxiety, abnormal eye movements, and constipation. Seizures were also a major feature, occurring in 48% of individuals. Interestingly, in our cohort, while we observe differences between sexes, particularly in areas such as coordination, unusual movements, obesity, impulsivity, and repetitive stereotypic behavior, many features were present in both males and females at similar rates (Supplemental Figures 1-2). However, when considered in the broader context of all individuals described in the KDM5C literature, differences between sexes may become clearer. This was seen in our meta-analysis of seizures and intellectual disability, in which a larger proportion of males (and a smaller proportion of females) displayed seizures and intellectual disability when compared to the RARE-X data set (Figures 2-3, Supplemental Figure 3).

The majority of the RARE-X variants described here are unique, adding to the large number of orphan alleles associated with KDM5C-NDD. We do, however, note several unrelated individuals/families with the same variant within our RARE-X study, between our study and previous reports, and between previous reports. These include Q199* (2 families), V504M (2 families),^7,31^ P531L (2 families),^39^ R599C (3 families),^7,21,39^ R694* (2 families),^36,43^ R943* (2 families)^55^ (Table S2). As additional participants with variants across the different domains of KDM5C are more thoroughly characterized and more families with overlapping variants are profiled, genotype-phenotype relationships may begin to be elucidated; however, a lack of standardized reporting and a limited number of individuals preclude this possibility at this time.

As we gain much-needed information regarding the spectrum of KDM5C-NDD-associated symptoms and their impact on daily life, we hope this knowledge will guide healthcare professionals, direct service providers, and others with limited exposure to this rare disorder in recognizing the potential features that may be present or emerge. Phenotypic data provided here is also likely to assist in the development of screening and assessment tools to guide clinical management of KDM5C-NDD and identify areas for additional intervention or testing, similar to resources available for other genetic conditions and NDDs.^95–99^ Given the age range of participants (2-20 years) and the overall communication challenges noted in participating individuals, it is likely that our study does not provide an all-inclusive picture of the symptoms associated with KDM5C-NDD. Some symptoms may not be present yet or may not be able to be effectively conveyed to the caregiver completing the survey. Because RARE-X prompts participants to update their survey responses yearly, going forward, we expect to gain additional insights into how the changing needs and presentation of older adults may differ from those of the children and young adults studied thus far. In the future, increasing recruitment efforts aimed at older participants or other underrepresented groups, such as international participants, will enhance the representation of these groups in our data. The ongoing nature of the RARE-X data collection also enables us to increase the total number of participants in our study and address the participant drop-off typically seen with more level 2 surveys that provide detailed information about specific topics in the future.

Based on the limited understanding of the association between pathogenic variants in *KDM5C* and epilepsy, we developed a *Drosophila* model of seizure-like behavior to enable molecular investigations of this association. To enable robust investigations, we optimized three different paradigms to quantify seizure-like behavior that utilize a new 3D-printed vial holding apparatus (Figures 4). More importantly, our study demonstrates that KDM5 is crucial for normal neuron function, as its loss in neurons leads to seizure-like behavior (Figure 5). This was a feature not seen when KDM5 was reduced in glia or in the mushroom body, despite altered axonal growth and guidance of mushroom body neurons being a characteristic of *Kdm5* knockdown flies (Figures 5-7). Future refinement of the types of neurons that contribute to this phenotype, as well as the brain regions that function as seizure epicenters, will reveal important information regarding KDM5-related seizures. Transcriptomic and cellular assays will also provide opportunities to define the signaling pathways that are perturbed and lead to seizure susceptibility. This, in turn, is likely to highlight pathways for the development of targeted therapeutics. The scalability of *Drosophila* studies provides a unique opportunity to conduct large-scale screens to test the efficacy of new and existing anti-epileptic medications using our knockdown animals, as has been done previously with other *Drosophila* epilepsy models.^100,101^

Extending our studies using *Drosophila*, we anticipate in the future including studies of seizure susceptibility with age, as this animal model has a median lifespan of approximately 50 days. The studies described here focused on younger adult flies (0-7 days in age). Other seizure models show changes with age, indicating that older flies may exhibit more dramatic differences in seizure-like behavior susceptibility than young flies.^102,103^ *Drosophila* larvae have also been used to model seizures, and analyzing this developmental time point may help identify critical windows for KDM5 function in preventing seizures.^77^ Like the intersection between age and seizures, we could investigate the interactions between seizures and other traits, such as altered sleep or diet/microbiome, as has been studied in other fly models.^104–108^

Taken together, our human and *Drosophila* findings underscore the critical role that KDM5 proteins play in nervous system functioning, where their loss or dysfunction predisposes to altered cognition and seizures/seizure-like behavior. By capturing the seizure and other behavioral features of KDM5C-NDD, our *Drosophila* model provides a means to understand the basis of traits observed in humans. Studying more cases of individuals with variants in *KDM5A*, *KDM5B*, and *KDM5C*, along with modeling these alleles in *Drosophila*, will broaden our understanding of the mechanisms underlying KDM5 family-related neurodevelopmental conditions.

## IV. Materials and Methods

### Subjects surveyed through RARE-X

All research involving human participants was conducted with approval from the Albert Einstein College of Medicine Institutional Review Board (IRB) (protocol number #2024-15912). Access to data collected by RARE-X, a program of Global Genes, was obtained under a Data Use Agreement between Global Genes and the Albert Einstein College of Medicine. RARE-X is a 501(c)(3) non-profit that collects and stores health-related information about patients with rare diseases. Informed consent was obtained by Global Genes/RARE-X for each participant/caregiver.

For simplicity, we refer to all individuals described in this study as “participants” or “respondents” when referring to information gathered by RARE-X-administered surveys, regardless of whether responses were provided directly by the individual with a *KDM5C* variant or by a caregiver proxy. Any individual with a *KDM5C* variant could enroll (directly or by proxy) in the KDM5C Data Collection Program by requesting access to the RARE-X platform. Inclusion criteria for this study included any person who has a confirmed variant in the *KDM5C* and has uploaded a genetic report for validation by RARE-X. Those without curatable genetic reports were excluded from the current analysis. For most participants, the variants reported were described in reference to isoform 1, the longest isoform, of *KDM5C*. Two individuals (Xp11.22 (53242376_53247160) and H514L) did not have a RefSeq accession number provided but were assumed to be referencing isoform 1. One participant was originally described in reference to isoform 2 of KDM5. For comparison purposes, this variant was realigned to isoform 1, changing the reported variant from R1185C (isoform 2) to R1252C (isoform 1).

For this study, the cohort of 31 participants comprised of those who have provided curatable genetic reports between March 2023 through June 2025. Participants represented 5 countries across the globe, with predominant representation from the United States (25), and representation from United Kingdom (3), Canada (1), Mexico (1), Germany (1). Enrolled individuals (or their caregivers if applicable) were given several initial surveys to collect basic, demographic, and overall health information and were asked to upload their genetic reports for curation and de-identification by the RARE-X team. Of these initial surveys, one particularly important survey included the Health and Development-Head-to-Toe survey, where participants were asked to note whether a certain area or function of the body was affected with a “Yes”, “No”, or “Unsure”. Based on responses to the Health and Development-Head-to-Toe survey, level 2 (L2) surveys were assigned by RARE-X to gain a more comprehensive picture of the symptoms observed in each participating individual. Fewer responses were noted as survey level increased, in part because non-affected individuals should not have been assigned L2 surveys to complete.

Firstly, three participants with the same confirmed genetic variant (c.4118-9A>G) from the same country were enrolled in this study. Given that two of the three report familial/targeted variant testing that was done the following year after the first participant’s whole exome sequencing and without any features excluding the possibility that these three individuals are related, they were treated as one family. Two different individuals presented with the same variant (Q199*), however these were ruled as unrelated individuals as they were from different countries and the female participant is noted as having a *de novo* variant.

Three individuals report variants that have been previously noted in the literature (L257Afs*5, P531L and R943*). For L257Afs*5, due to age (with our participant being much younger), reporting that the female participant is unsure about a family history, and the female participant being from different country in the same continent than the where the literature study was conducted, this participant is being treated as a unique individual from a different family.^109^ For P531L, the individuals were different sexes from different continents and thus treated as unique individuals from different families.^39^ For R943*, the individuals are from different continents and both report *de novo* inheritance and were thus treated as unique individuals from different families.^55^

### Literature Review

A literature search was conducted using different combinations of a variety of search terms, some of which included “Claes-Jensen Syndrome”, “KDM5C”, “JARID1C”, “SCMX”, “X-linked intellectual disability”, “neurodevelopmental”, and “developmental”. Identified journal articles were reviewed for the mention of intellectual disability, intellectual disability level, seizures, seizure type, seizure-related medications taken, protein/genetic variant, sex, and inheritance type. If a variant was reported to be inherited, the parent from whom it was inherited from (if described) was included as a *KDM5C*-variant possessing individual in the literature review table, even if no additional information was included (Table S2). If sex or inheritance type was not explicitly mentioned, it was reported as “Unknown”.

When assessing intellectual disability and seizure features at an individual level, if a feature (Seizures/Intellectual Disability) was explicitly noted to be present, the feature was marked as a “Yes”. If a feature was explicitly noted to not be present, the feature was marked as a “No”. If a feature was not explicitly mentioned, the feature was reported as “Not Described/Not Assessed” or “(ND/NA)”. When assessing intellectual disability and seizure features at a family level, families were marked as “Yes” if any member of the family reported to have that feature. Family-level features as “No” if it was explicitly stated that none of the family members have that feature. Family-level features were marked as “No/ND/NA” if there were no reports of that feature, but not every single family member was explicitly defined as to not have that feature.

#### Fly strains and care

Flies used in this study were kept at 25°C on a 12-hour light:dark cycle. A standard cornmeal/molasses/yeast diet was provided.^68^ Male and female flies were analyzed for each experiment, and data were pooled if no sex-based difference was detected. The Elav-Gal4 strain used (see below) also contained tub-Gal80^ts^, but this was kept inactive by raising the animals at 25 °C, allowing knockdown in neurons.

The following fly strains were used in this study:

| Source | Nomenclature used | Genotype |
| --- | --- | --- |
| BDSC #602908 | *repo-Gal4* | w[1118]; P[w[+mC]=repo-Gal4.L]Oatp30B[repoF3] |
| BDSC #458 | *elav-Gal4* | P[GawB]elav<sup>C155</sup> |
| BDSC #7017 | *tub-Gal80<sup>ts</sup>* | P[tubP-Gal80<sup>ts</sup>]2 |
| BDSC #854 | *OK107-Gal4* | w[*]; P[w[+mW.hs]=GawB]OK107 ey[OK107]/In(4)ci[D], ci[D] pan[ciD] sv[spa-pol] |
| BDSC #35706 | *UAS-shKdm5* | y[1] sc[*] v[1] sev[21]; P[y[+t7.7] v[+t1.8]=TRiP.GLV21071]attP2/TM3, Sb[1] |
| Navarro-Costa et al., 2016<sup>110</sup> | *Kdm5:HA* | *Kdm5<sup>Δ</sup>* ; *gKdm5:HA* (attP86F) |
| Hatch et al., 2021<sup>69</sup> | *Kdm5:3xHA* | *Kdm5:HA* (endogenous knock-in) |

#### Seizure induction techniques

For all seizure tests, fly vials were plugged with Charlie Plugs (Jaece CP-N730) that were cut at an angle to ensure that no flies were occluded from view during testing. Flies were tested at an age of ≤ 7 days post-eclosion, and the sex of the flies was noted for each vial/trial. The day before intended testing, flies were separated into food-containing vials in groups of ≤7 flies and allowed to recover overnight after being anesthetized with carbon dioxide. During the day of testing, flies were transferred into empty vials and allowed to recover for at least an hour before they were tested. Vials were recorded with an iPhone or iPad mini camera, and all testing was conducted between 9am and noon.

Mechanical seizures were induced by individually shaking vials on a laboratory vortex (Fisher Scientific #02215365) at maximum speed for ten seconds at room temperature. Immediately post-vortexing, vials were moved to custom designed vial holders (see 3D Printing methods section for more information). Vials were recorded for one or five minutes after shaking. This protocol was adapted from Mituzaite et al., 2021.^77^ Heat seizures were induced by placing fly vials directly into a 40°C water bath for five minutes. A twelve-quart sous vide container (EVERIE Sous Vide Container Bundle from Amazon.com) was used as a water bath and a consistent water temperature was maintained by an immersion circulator originally meant for Sous Vide cooking (Anova Culinary Nano Sous Vide Precision Cooke, #AN400-US00). Plugs were pushed down to around the 75% of the height of the vial so that the vials could be submerged to a point where the area in which the flies could move would be completely covered by the water in the water bath, but not so much that water could enter in through the top. Vials were held in the water with a custom designed vial holder (see 3D Printing methods section for more information) that was weighed down by a brick (8” x 4” x 2”, Lowe’s #817669) or a four-way modular tube rack (Cole-Parmer UX-06733-00). Flies were recorded for five minutes post water bath exposure for later analysis. This protocol was adapted from Mituzaite et al., 2021.^77^

Spontaneous seizures were defined as any seizure-like activity during a five-minute period of time without any stressor (heat or mechanical) given.

#### Behavioral analysis

Videos were assessed in Adobe Premiere Pro (v.24.5.0) by adding markers to video clips when a fly displayed seizure-like activity. The markers were added so that the “In” time was the beginning of the behavior and the “Out” time was the end of that behavior. This was done for each time that an individual fly seized, so that the following parameters could be measured: seizure presence (yes/no), total seizure duration, time to first seizure, and number of seizures. Seizure susceptibility and duration was based on the period during which a fly turned on its side or back and displayed shaking or immobilization (paralysis). This is a characteristic part of the *Drosophila* seizure cycle and the clearest visual to determine if a fly demonstrates seizure-like behavior.^74^ An event was defined as seizure-like behavior for all events lasting at least one second.

#### 3D printing

For video recording purposes, flies were held in either a four-way modular tube rack (Cole-Parmer UX-06733-00) with a white backdrop or one of two custom-designed 3D-printed vial holder racks. The vial holders were designed in Tinkercad (Autodesk, https://www.tinkercad.com/) and printed in white Polylactic Acid (PLA, Flashforge) using an Adventurer 4 Pro 3D printer (Flashforge). The larger vial holder was custom designed with water bath heat-induced seizure assays in mind and was created to accommodate an immersion circulator and a brick or some other sort of weight. The smaller vial holder was designed with mechanical seizure induction in mind and created to be used out of water. STL files are available as supplemental files.

#### Antibodies

The following antibodies were used in this study:

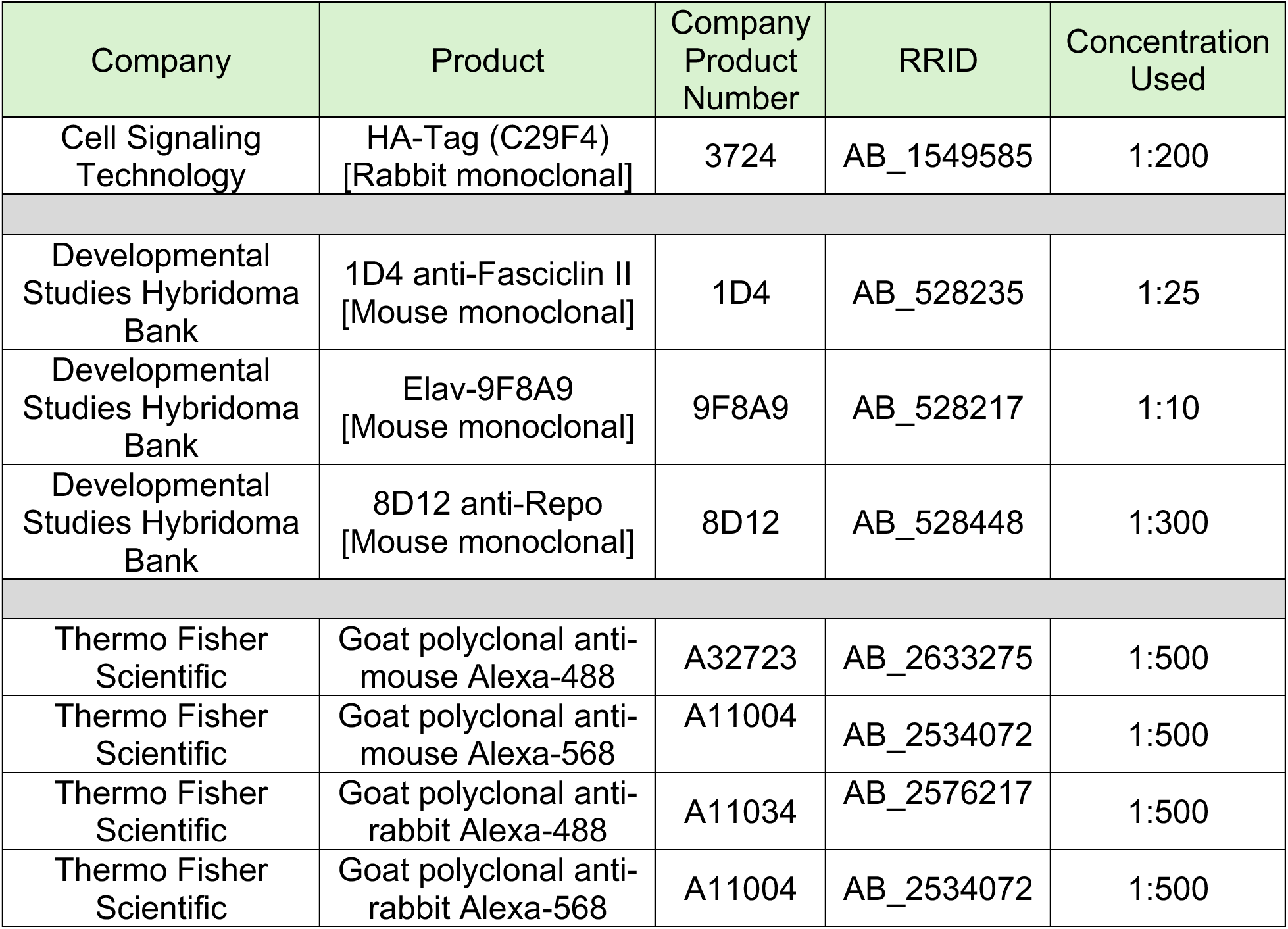

| Company | Product | Company Product Number | RRID | Concentration Used |
| --- | --- | --- | --- | --- |
| Cell Signaling Technology | HA-Tag (C29F4)<br>[Rabbit monoclonal] | 3724 | AB_1549585 | 1:200 |
| Developmental Studies Hybridoma Bank | 1D4 anti-Fasciclin II<br>[Mouse monoclonal] | 1D4 | AB_528235 | 1:25 |
| Developmental Studies Hybridoma Bank | Elav-9F8A9<br>[Mouse monoclonal] | 9F8A9 | AB_528217 | 1:10 |
| Developmental Studies Hybridoma Bank | 8D12 anti-Repo<br>[Mouse monoclonal] | 8D12 | AB_528448 | 1:300 |
| Thermo Fisher Scientific | Goat polyclonal anti-mouse Alexa-488 | A32723 | AB_2633275 | 1:500 |
| Thermo Fisher Scientific | Goat polyclonal anti-mouse Alexa-568 | A11004 | AB_2534072 | 1:500 |
| Thermo Fisher Scientific | Goat polyclonal anti-rabbit Alexa-488 | A11034 | AB_2576217 | 1:500 |
| Thermo Fisher Scientific | Goat polyclonal anti-rabbit Alexa-568 | A11004 | AB_2534072 | 1:500 |

#### Immunohistochemistry

Adult *Drosophila* of ≤7 days of age were fixed in 4% paraformaldehyde (PFA, Thermo Scientific™ #28908) in phosphate buffered saline (PBS) + 0.2% Triton X-100 (Fisher Scientific #BP151-100) (PBS-T) for three hours. Fixed flies were washed three times for fifteen minutes each with PBS-T at room temperature. Whole brains were dissected out and then blocked for an hour in a solution of 4% normal donkey serum (NDS, LAMPIRE Biological Laboratory # 7332100) in PBS-T. Brains were then incubated overnight in primary antibody solution consisting of primary antibodies, 4% NDS, and PBS-T. The following day brains were washed three times for fifteen minutes with PBS-T before incubation overnight in secondary antibody solution consisting of secondary antibodies, 4% NDS, and PBS-T. After one more set of three fifteen minutes washes in PBS-T the next day, brains were mounted onto slides in DAPI Fluoromount-G (SouthernBiotech™ #010020).

#### Image Acquisition

Representative images were taken on a Leica SP8 confocal microscope with a 40x oil immersion lens. Images were taken at either a size of 1024 x1024 pixels with a speed of 400 hz or a size of 1024 x 1024 pixels with a speed of 100 hz. Image z-stacks taken to assess mushroom body morphology were acquired with a step size of 1 µm. Image stacks were processed in the Leica Application Suite X software program (Leica 3.7.6.25997). Figure illustrations were created with BioRender.com and Adobe Illustrator (v.29.8.1). Figures were assembled in Adobe Photoshop (v. 25.9.1).

#### Statistical Analysis

Statistical analyses (including Fisher’s exact tests, unpaired two-tailed Mann-Whitney tests, unpaired two-tailed t-tests, and Log-rank (Mantel-Cox) tests) were conducted using GraphPad Prism (v.10.2.2). For the RARE-X related studies and the combined literature and participant meta-analyses, the participant number (broken down by sex) are reported in the figure legend and statistical analyses (if completed) were included in the figure and/or figure legend for each analysis conducted. For the *Drosophila* studies, animal number evaluated, statistical analyses, and p-values are noted in each figure or legend for each analysis conducted.

## Data Availability

NA

## Acknowledgements

Thank you to the KDM5C families who participated in this study – we are incredibly appreciative of your time and insights. We are also grateful to Melissa Murr McNeilly, Mariam Rebollar, and the entire KDM5C Advocacy, Research, Education & Support (KARES) Foundation for their ongoing efforts to support the KDM5C Data Collection Program. Survey and genetic curation data utilized in this publication was provided by RARE-X, a research program of Global Genes. RARE-X provides a collaborative platform for global data sharing and analysis to accelerate research for rare diseases. Global Genes is a 501(c)(3) non-profit organization dedicated to eliminating the burdens and challenges of rare diseases for patients and families globally. We would like to thank Geoffrey Beek and Dr. Zohreh Talebizadeh for their consultation and assistance and the entire RARE-X team for maintaining and curating the provided data.

We thank members of the Secombe lab for their feedback. We are also appreciative of Hillary Guzik and the Einstein Analytical Imaging Facility (AIF) for training and access to the Leica SP8 Confocal Microscope (Shared Instrumentation Grant 1S10OD023591-01). Stocks obtained from the Bloomington Drosophila Stock Center (NIH P40OD018537) were used in this study. Antibodies obtained from the Developmental Studies Hybridoma Bank, created by the NICHD of the National Institutes of Health (NIH) and maintained at The University of Iowa, were also used in this study. This work was supported by the NIH (B.K.T was supported by NIGMS K12GM102779, A.M. was supported by T32GM149364, and J.S. by P50HD105352 & R01GM150189).

## V. Author Contributions

Conceptualization: BKT, JS

Methodology: BKT, AM, BL

Data curation: AM, BKT

Formal Analysis and Visualization: BKT

Investigation: BKT, AM, BL, JS

Formal Analysis and Visualization: BKT

Supervision: JS

Writing – original draft: BKT, JS

## VI. Declaration of Interests

Julie Secombe is the Chair of the Scientific Advisory Board for the KDM5C Advocacy, Research, Education, & Support (KARES) Foundation (volunteer position).

---

**Supplemental Figure 1.**
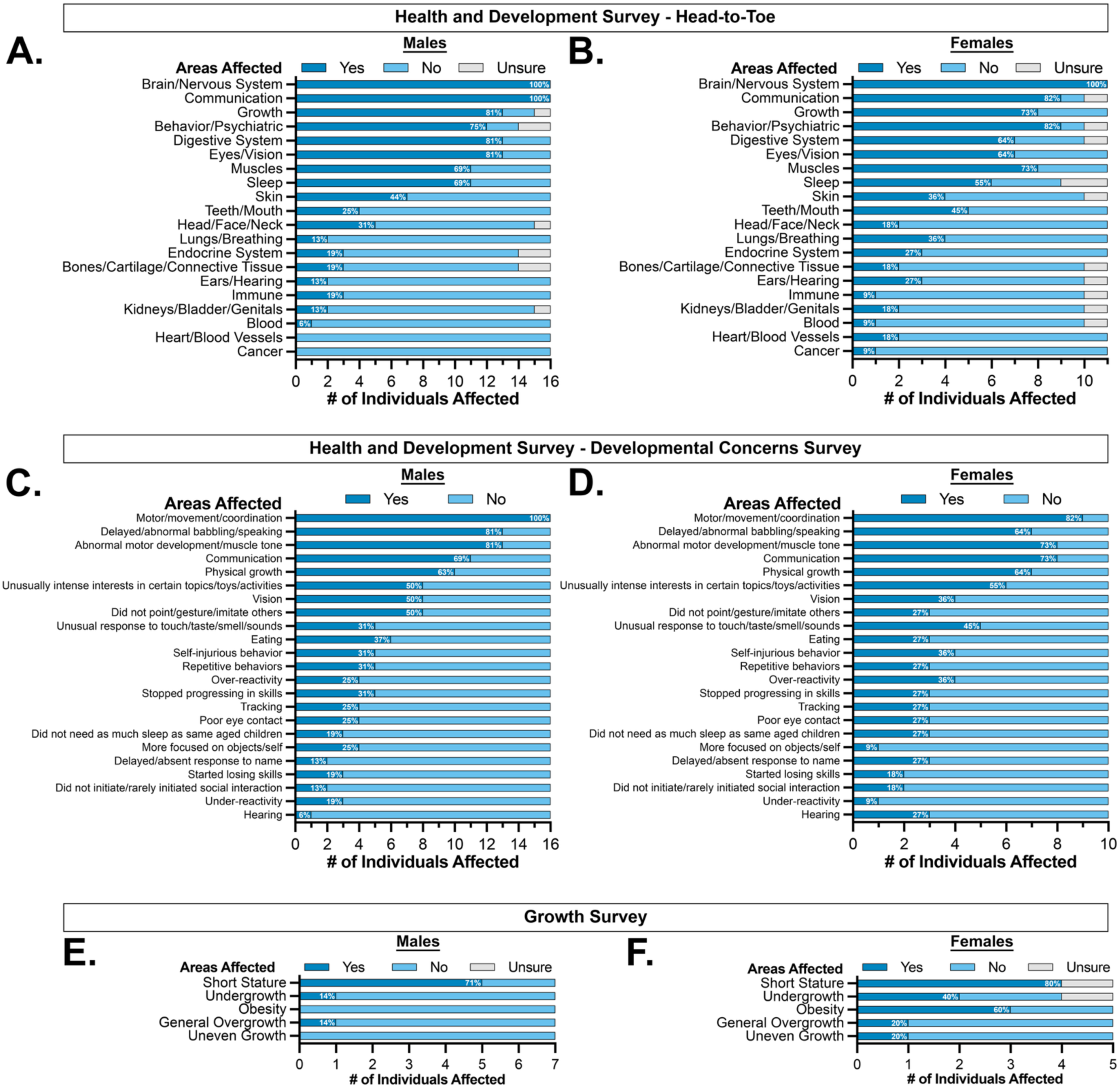
Male and female individuals from the RARE-X cohort with KDM5C variants note overlapping, but not identical, issues across the body. (A-B) Assessment of completed Health and Development-Head-to-Toe surveys revealed that while all participants, regardless of sex, have reported nervous system issues, participants show a variety of concerns across multiple body systems. (C-D) Assessment of completed Health and Development-Developmental Concerns surveys showed that motor issues were the most frequently reported developmental concern for both male and female participants. Other similar concerns were noted between sexes but varied in prevalence. (E-F) Assessment of completed level 2 (L2) Growth surveys identified short-stature as the most common growth-related concern for both males and females. Other features, such as obesity and undergrowth, however, were seen in a greater proportion of female than male participants.

**Supplemental Figure 2.**
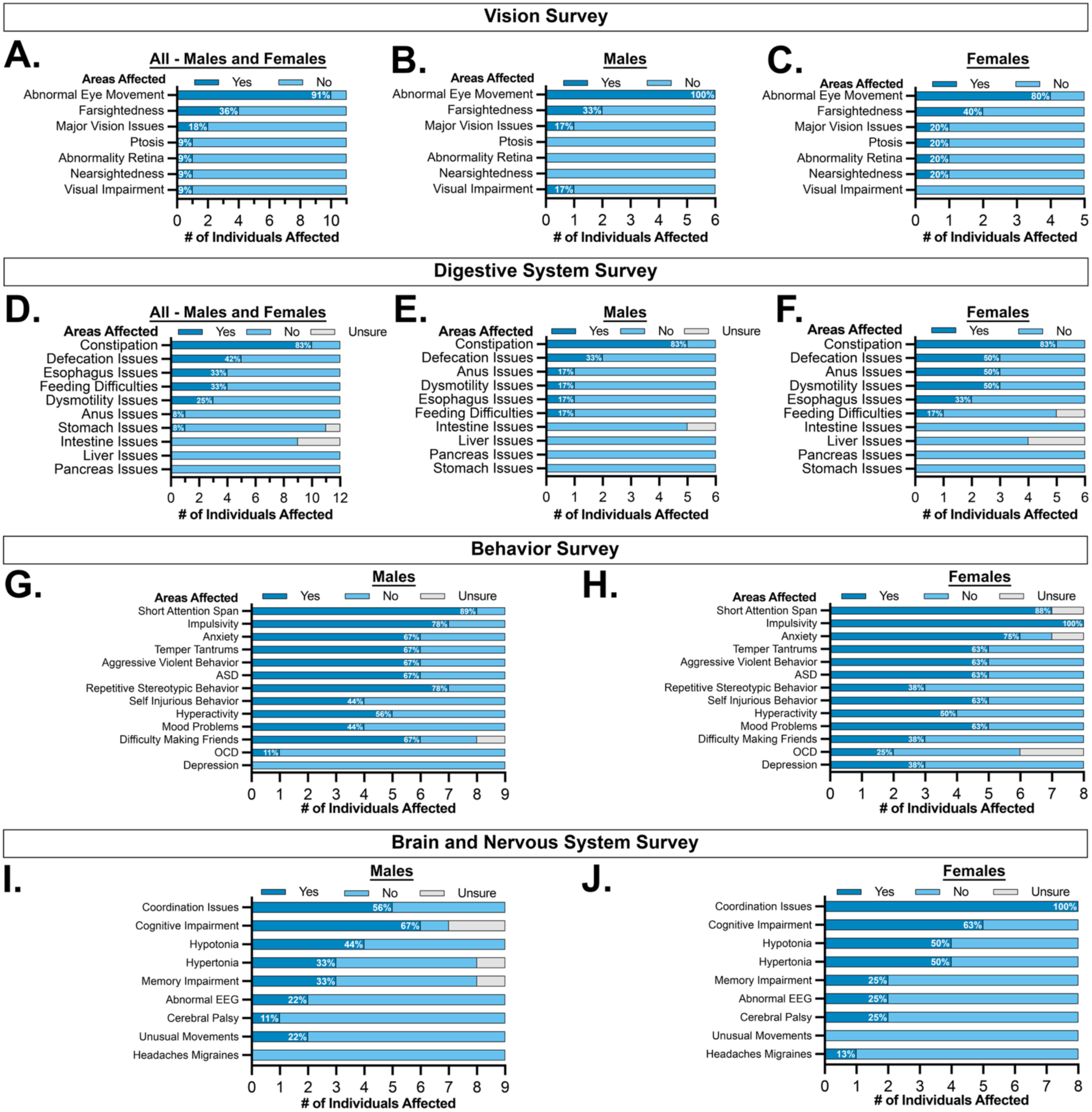
Male and female individuals with KDM5C variants from the RARE-X cohort are affected in the realm of behavior as well as in the realms of nervous, visual, and digestive system function. (A-B) Assessment of completed level 2 (L2) Vision surveys showed that the most common issues reported were abnormal eye movements and farsightedness, although other issues, which included major vision issues, ptosis, retina abnormalities, nearsightedness, and visual impairment, were noted. (C-D) Assessment of completed L2 Digestive System surveys revealed that the majority of participants, male and female, reported issues with constipation. However, other challenges, including issues with defecation, feeding, dysmotility, and the anus were reported. (E-G) Assessment of completed L2 Behavior surveys revealed that short attention span, impulsivity, and anxiety were some of the most frequently reported behavior-related concerns amongst male and female participants, however, multiple other behavioral issues were noted in both sexes. Abbreviations Used: Autism Spectrum Disorder (ASD), Obsessive Compulsive Disorder (OCD). (H-J) Assessment of completed L2 Brain/Nervous systems surveys indicated that male and female participants showed overlapping symptoms, however, several symptoms were seen in only males or females (ex. unusual movements and headaches/migraines. Abbreviations Used: Electroencephalography (EEG).

**Supplemental Figure 3.**
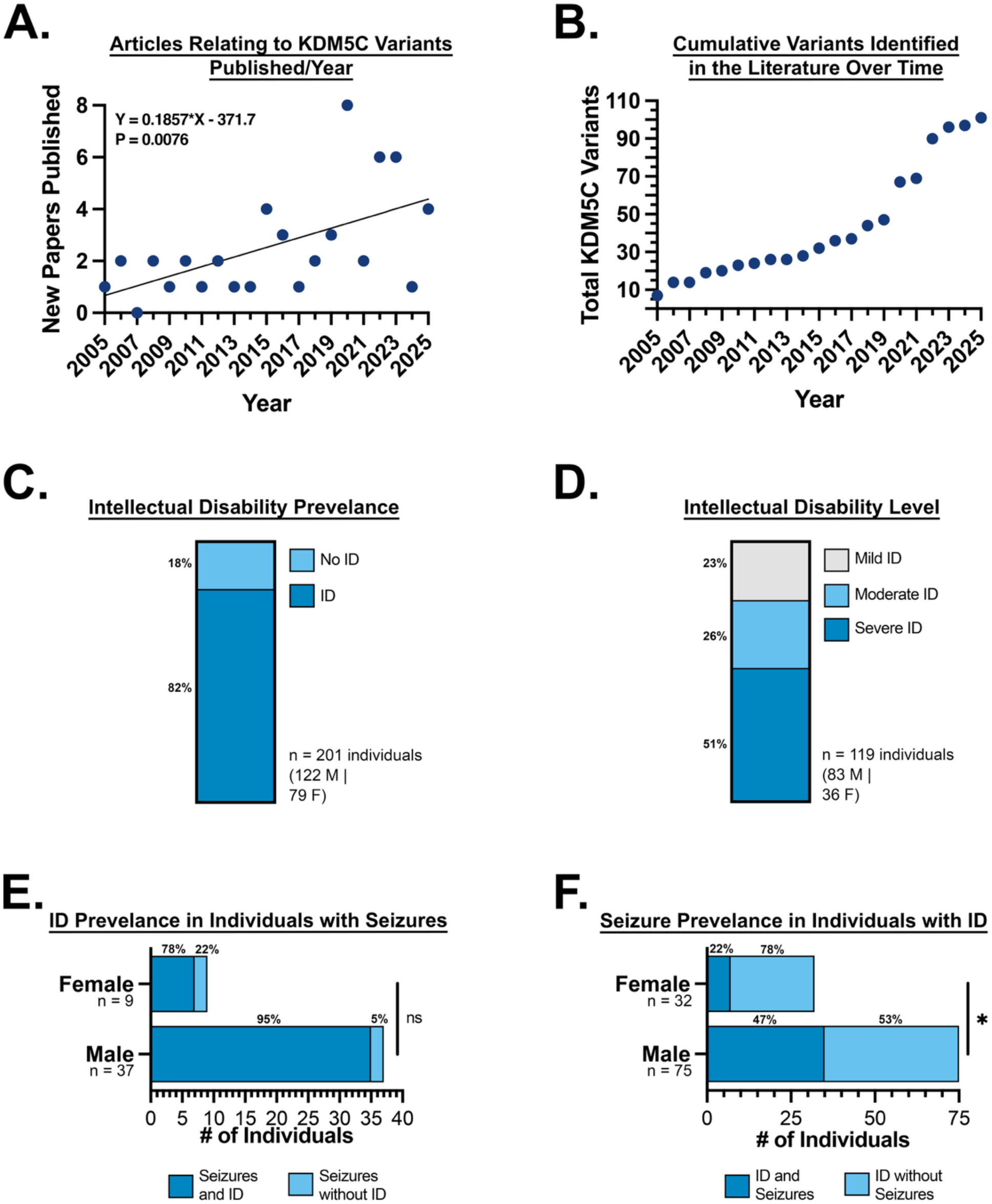
Seizures and intellectual disability are seen in male and female individuals in both previous reports and in the RARE-X cohort of individuals with KDM5C variants. (A) Visualization of the number of journal articles that detail KDM5C variants by year illustrates an increase in the number of articles published over time. Statistical analysis was done using a simple linear regression. (B) Visualization of the cumulative number of KDM5C variants identified in the literatures shows that new variants have been added in most years since the first article in 2005. A total of 101 variants have been identified in the literature (at the time of article submission). (C) Analysis of the data collected by RARE-X and of information from previously published works demonstrates that intellectual disability (ID) is seen in over three-quarters of individuals with KDM5C variants. (D) Analysis of information from previously published works demonstrates that intellectual disability (ID) levels vary in individuals with KDM5C variants, with over half of participants exhibiting severe ID, around 26% exhibiting moderate ID, and 23% exhibiting mild ID. (E) Analysis of the data collected by RARE-X and of information from previously published works shows that males with seizures are more likely to also present with ID than females with seizures. Statistical analysis was done using a Fisher’s Exact test (two-sided, P = 0.1667). (F) Analysis of the data collected by RARE-X and of information from previously published works shows that males with ID are more likely to also present with seizures than females with ID. Statistical analysis was done using a Fisher’s Exact test (two-sided, P = 0.0183). *P < 0.05.

**Supplemental Figure 4.**
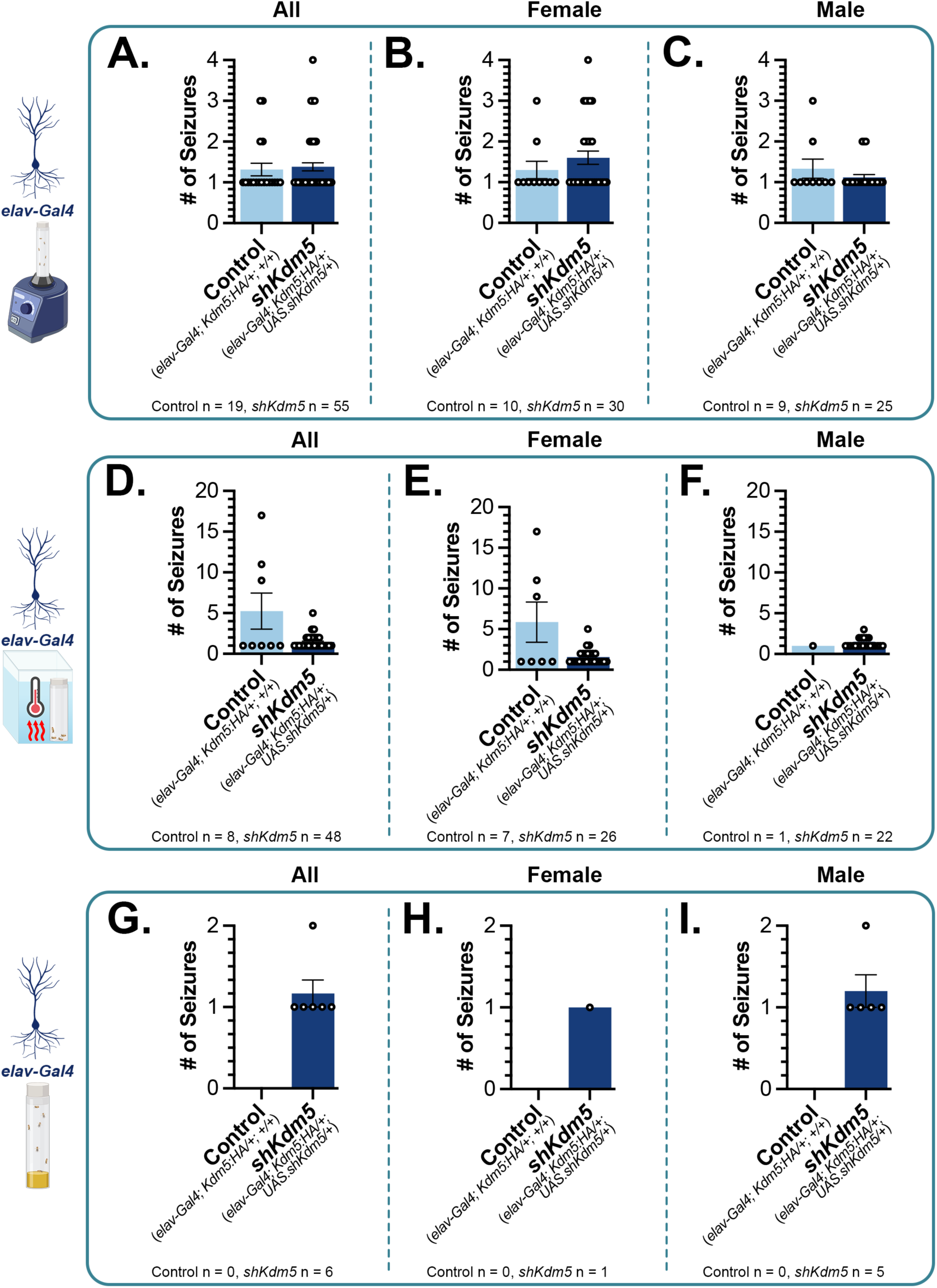
Number of seizure events does not differ between *Kdm5* knockdown flies and controls. (A-C) For flies that seized following mechanical stress, the number of seizure-like events did not significantly differ between *elav*>+ control (*elav-Gal4*; *Kdm5:HA/+*; *+/+*) and *elav*>*shKdm5* knockdown (*elav-Gal4*; *Kdm5:HA/+*; *UAS-shKdm5/+*) groups when the data was pooled or separated by sex. Statistical analyses were done using a Fisher’s Exact test (two-sided, P = 0.7422 for panel A, P = 0.3778 for panel B, P = 0.4123 for panel C). (D-F) Following heat stress, although far fewer flies seized overall in the control group, *elav*>+ control flies had more seizure-like events than *elav*>*shKdm5* knockdown flies when pooled or when the female flies only were examined. Statistical analyses were done using a Fisher’s Exact test (two-sided, P = 0.3592 for panel D, P = 0.322). Statistical analysis for panels F was not able to be completed because only one control fly seized. (G-I) Without a provoking stimulus, no *elav*>+ control flies had seizure-like events. However, *elav*>*shKdm5* knockdown flies had between 1-2 events. Statistical analysis for panels G-I were not able to be completed because no control flies seized.

**Supplemental Figure 5.**
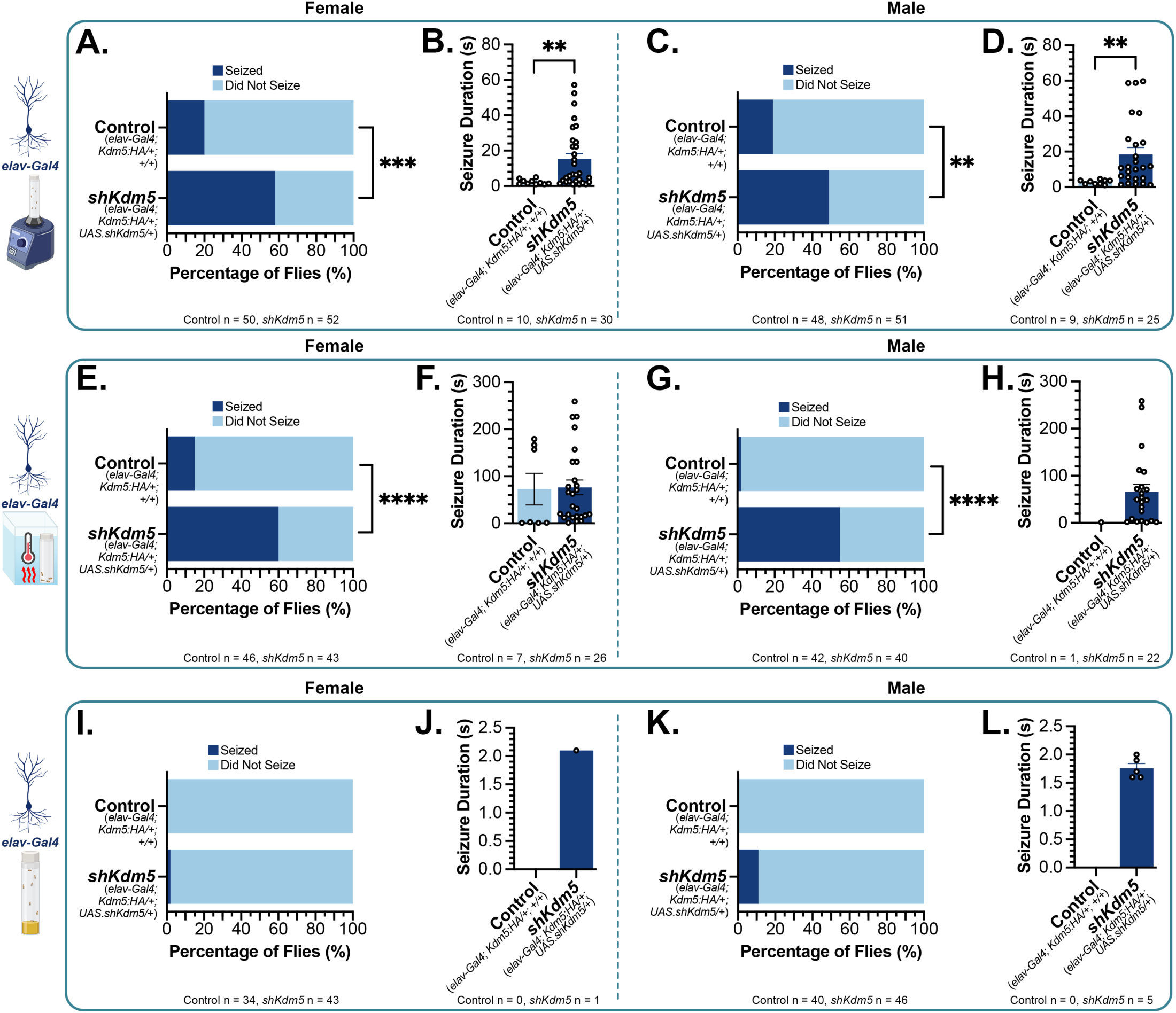
Both male and female flies seize more frequently, and for longer periods, than controls following KDM5 reduction in neurons. (A-D) Both male and female *elav*>sh*Kdm5* knockdown (*elav-Gal4; Kdm5:HA/+; UAS-shKdm5/+*) flies seized significantly more frequently and for longer durations than *elav*>+ controls (*elav-Gal4; Kdm5:HA/+; +/+*) in response to mechanical stress. Statistical analysis for panel A was done using a Fisher’s exact test (two-sided, P = 0.0001). Statistical analysis for panel B was done using a Mann-Whitney test (two-tailed, P = 0.0038). Statistical analysis for panel C was done using a Fisher’s exact test (two-sided, P = 0.0028). Statistical analysis for panel D was done using a Mann-Whitney test (two-tailed, P = 0.0011). ***P < 0.001, ****P < 0.0001. (E-H) Both male and female *elav*>*shKdm5* flies seized significantly more frequently, but not statistically longer, than controls in response to heat stress. Statistical analysis for panel E was done using a Fisher’s exact test (two-sided, P < 0.0001). Statistical analysis for panel F was done using a Mann-Whitney test (two-tailed, p = 0.3405). Statistical analysis for panel G was done using a Fisher’s exact test (two-sided, P < 0.0001). Statistical analysis for panel H was not able to be completed because no control flies seized. ****P < 0.0001. (I-L) When separated into male and female groups, *elav*>*shKdm5* flies are not statistically significantly different than controls in their seizure susceptibility or duration when observed without an induction stimulus (spontaneous seizures). Statistical analysis for panel I was done using a Fisher’s exact test (two-sided, P > 0.9999). Statistical analysis for panel J was not able to be completed because no control flies seized. Statistical analysis for panel K was done using a Fisher’s exact test (two-sided, P = 0.0583). Statistical analysis for panel L was not able to be completed because no control flies seized.

**Supplemental Figure 6.**
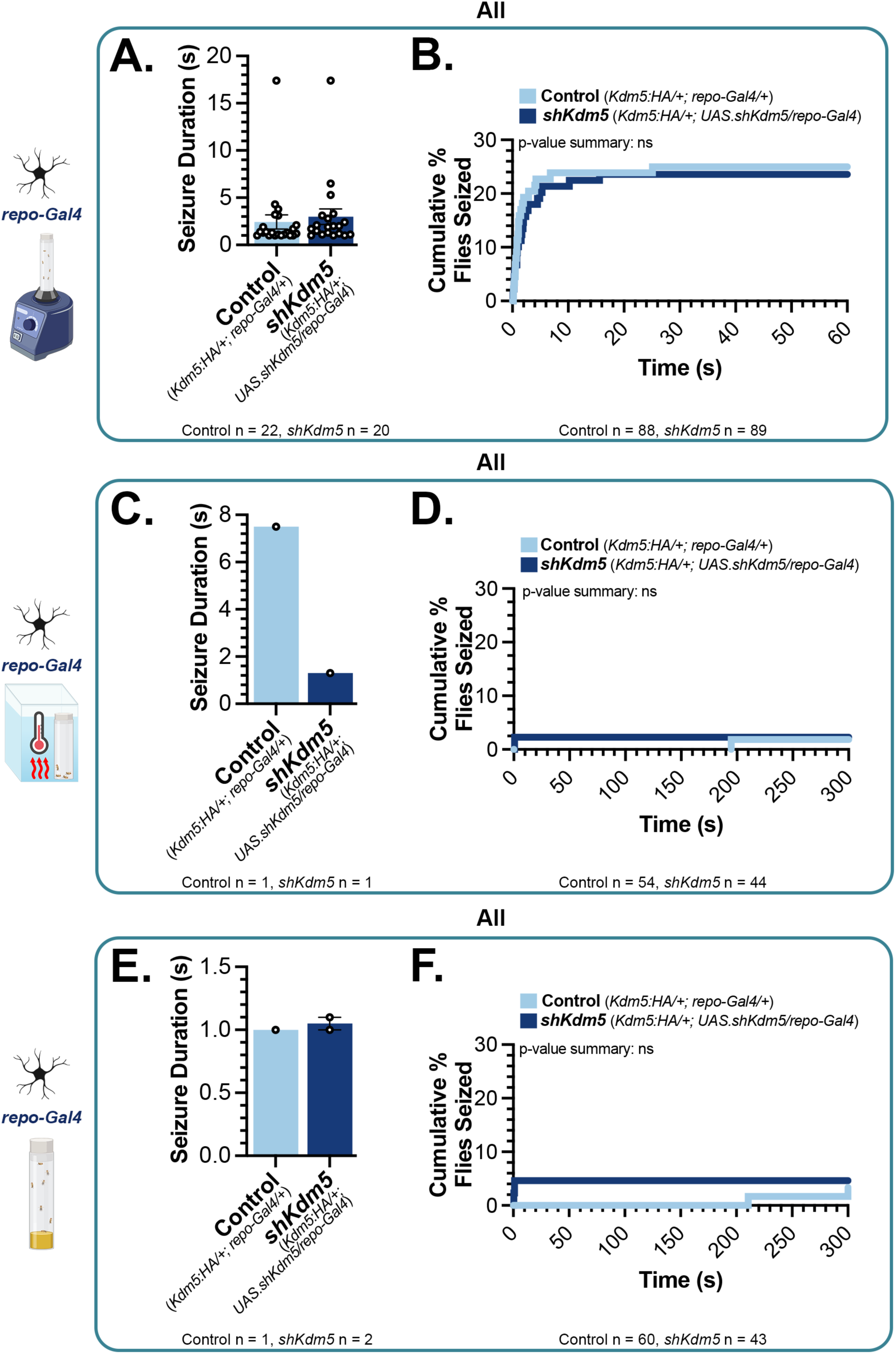
KDM5 reduction in glia does not increase seizure susceptibility or duration. (A-B) *repo>shKdm5* (*Kdm5:HA/+; UAS.shKdm5/repo-Gal4*) and *repo>+* (*Kdm5:HA/+; repo-Gal4/+*) control flies did not seize at statistically different rates following mechanical seizure induction. Statistical analysis for panel A was done using a Mann-Whitney test (two-tailed, p = 0.2246). Statistical analysis for panel B was done using a Log-rank (Mantel-Cox) test (P = 0.7943). (C-D *repo>shKdm5* and *repo>+* control flies did not seize at statistically different rates following heat seizure induction. Statistical analysis for panel C was not able to be completed because of the limited number of flies that seized. Statistical analysis for panel D was done using a Log-rank (Mantel-Cox) test (P = 0.8782). (E-F) *repo>shKdm5* and *repo>+* control flies did not seize at statistically different rates when observed for spontaneous seizures. Statistical analysis for panel E was not able to be completed because of the limited number of flies that seized. Statistical analysis for panel F was done using a Log-rank (Mantel-Cox) test (P = 0.7184).

**Supplemental Figure 7.**
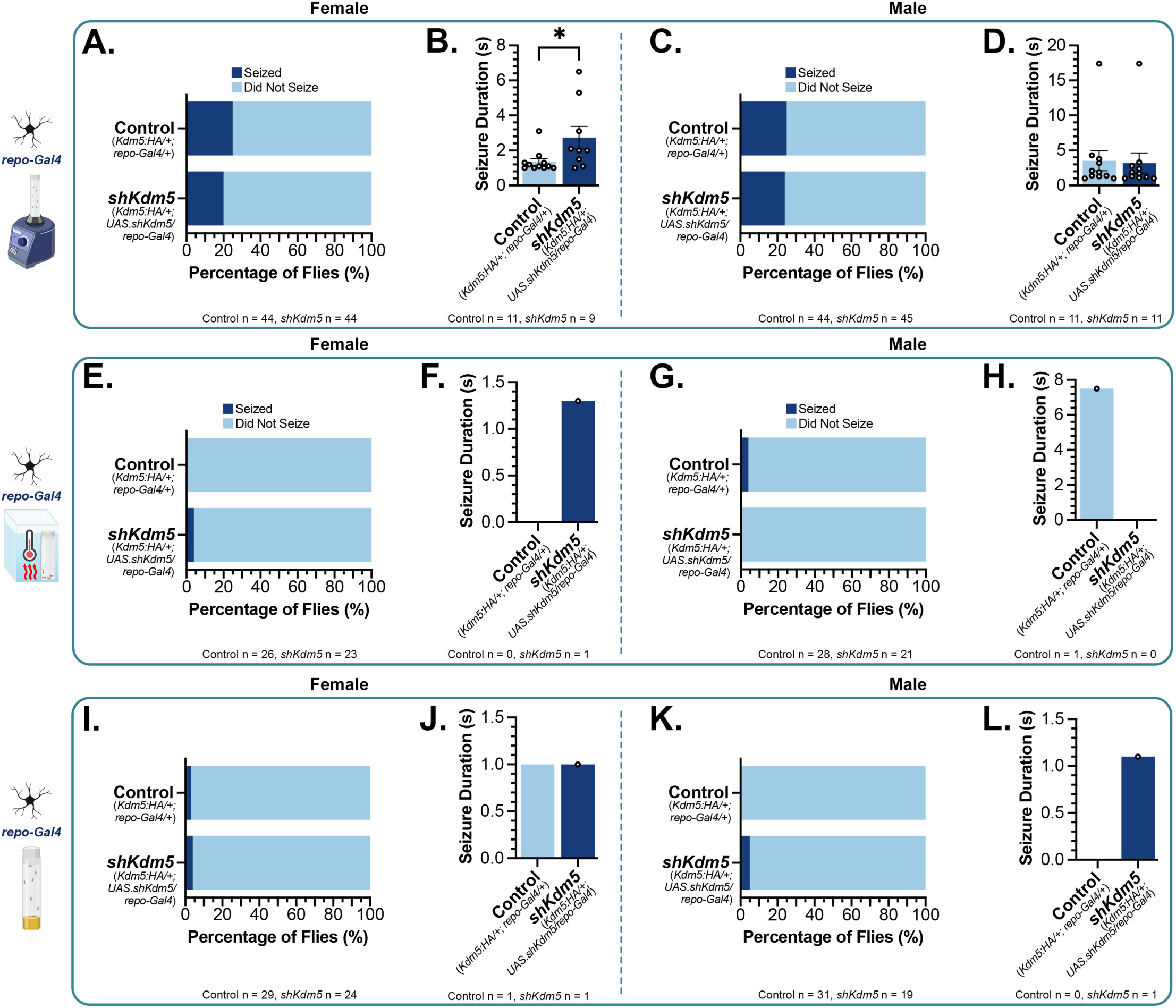
Neither male, nor female flies show an increased susceptibility to seize following KDM5 reduction pan-glially. (A-D) Following mechanical stress, *repo>shKdm5* (*Kdm5:HA/+; UAS.shKdm5/repo-Gal4*) and *repo>+* (*Kdm5:HA/+; repo-Gal4/+*) control flies differed only in seizure duration for female flies, but did not differ in terms of seizure susceptibility for female flies or seizure susceptibility/duration for male flies. Statistical analysis for panel A was done using a Fisher’s exact test (two-sided, P = 0.7997). Statistical analysis for panel B was done using a Mann-Whitney test (two-tailed, P = 0.0255). Statistical analysis for panel C was done using a Fisher’s exact test (two-sided, P > 0.9999). Statistical analysis for panel D was done using a Mann-Whitney test (two-tailed, P = 0.5711). (E-H) Neither male nor female *repo>shKdm5* differed from *repo>+* control flies in seizure susceptibility or duration following heat stress. Statistical analysis for panel E was done using a Fisher’s exact test (two-sided, P = 0.4694). Statistical analysis for panel F was not able to be completed because of the limited number of flies that seized. Statistical analysis for panel G was done using a Fisher’s exact test (two-sided, P > 0.9999). Statistical analysis for panel H was not able to be completed because of the limited number of flies that seized. (I-L) Neither male nor female *repo>shKdm5* differed from *repo>+* control flies in seizure susceptibility or duration when observed without a provoking stimulus (spontaneous seizures). Statistical analysis for panel I was done using a Fisher’s exact test (two-sided, P > 0.9999). Statistical analysis for panel J was not able to be completed because of the limited number of flies that seized. Statistical analysis for panel K was done using a Fisher’s exact test (two-sided, P = 0.3800). Statistical analysis for panel L was not able to be completed because of the limited number of flies that seized.

**Supplemental Figure 8.**
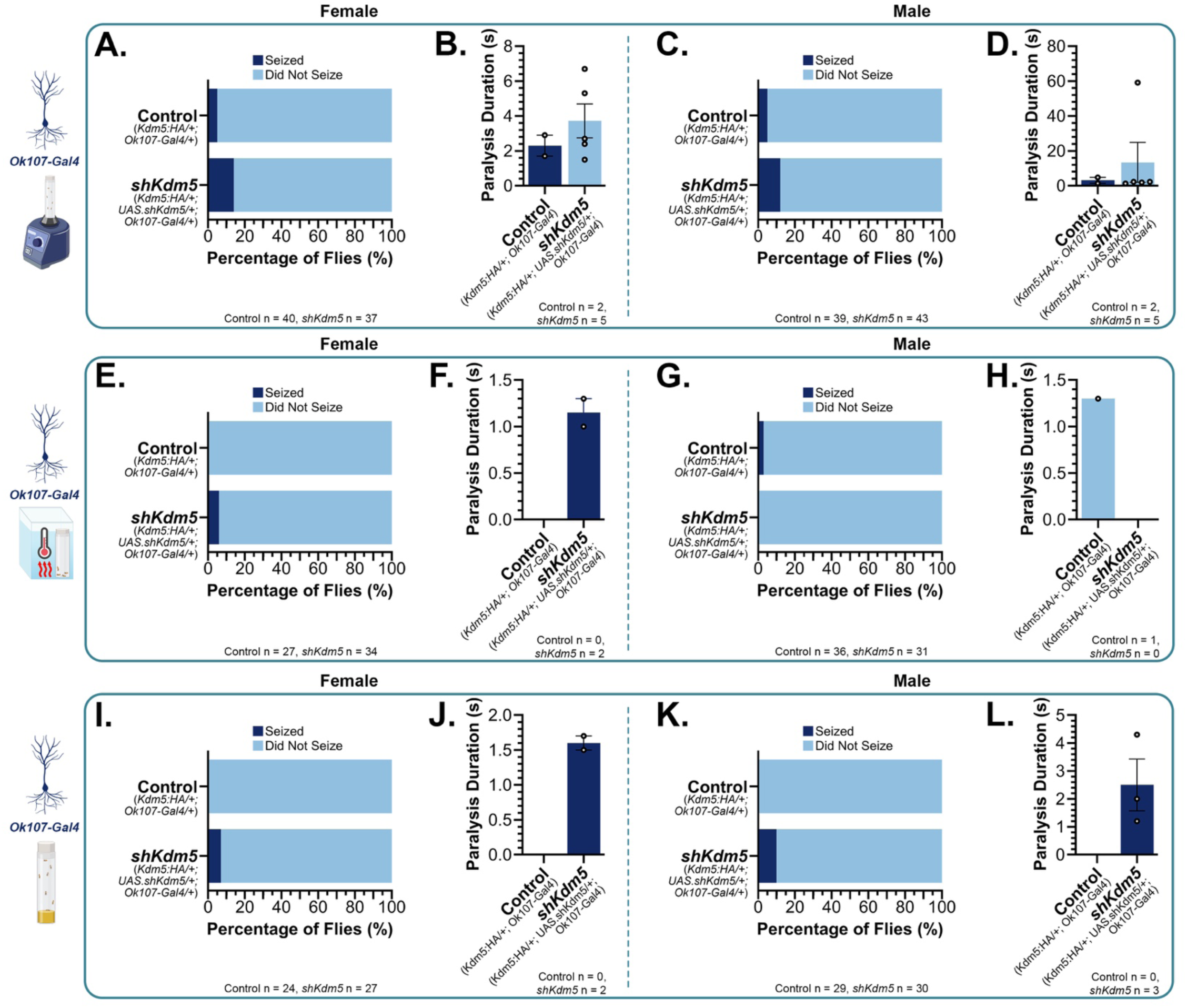
Neither male, nor female flies show an increased susceptibility to seize following KDM5 reduction in mushroom body neurons. (A-D) Neither male nor female knockdown *OK107*>*shKdm5* (*Kdm5:HA/+; UAS.shKdm5/+; OK107-Gal4/+*) flies differed from control *OK107*>+ (*Kdm5:HA/+; UAS.shKdm5/+; OK107-Gal4/+*) flies in seizure susceptibility or duration following mechanical stress. Statistical analysis for panel A was done using a Fisher’s exact test (two-sided, P = 0.2511). Statistical analysis for panel B was done using a Mann-Whitney test (two-tailed, P = 0.8571). Statistical analysis for panel C was done using a Fisher’s exact test (two-sided, P = 0.4363). Statistical analysis for panel D was done using a Mann-Whitney test (two-tailed, P > 0.9999). (E-H) Neither male nor female *OK107*>*shKDM5* flies differed from *OK107*>+ controls in seizure susceptibility or duration following heat stress. Statistical analysis for panel E was done using a Fisher’s exact test (two-sided, P = 0.4984). Statistical analysis for panel F was not able to be completed because of the limited number of flies that seized. Statistical analysis for panel G was done using a Fisher’s exact test (two-sided, P > 0.9999). Statistical analysis for panel H was not able to be completed because of the limited number of flies that seized. (I-L) Neither male nor female *OK107*>*shKDM5* flies differed from *OK107*>+ controls in seizure susceptibility or duration when observed without a provoking stimulus (spontaneous seizures). Statistical analysis for panel I was done using a Fisher’s exact test (two-sided, P = 0.4918). Statistical analysis for panel J was not able to be completed because of the limited number of flies that seized. Statistical analysis for panel K was done using a Fisher’s exact test (two-sided, P = 0.2373). Statistical analysis for panel L was not able to be completed because of the limited number of flies that seized.

